# Adenine base editing corrects point mutation in mitochondrial single-stranded binding protein (*SSBP1*) to improve mitochondrial function

**DOI:** 10.1101/2023.11.02.23297943

**Authors:** Ju Hyuen Cha, Seok-Hoon Lee, Yejin Yun, Won Hoon Choi, Hansol Koo, Sung Ho Jung, Ho Byung Chae, Dae Hee Lee, Seok Jae Lee, Dong Hyun Jo, Jeong Hun Kim, Jae-Jin Song, Jong-Hee Chae, Jun Ho Lee, Seung Ha Oh, Jiho Park, Jin Young Kang, Sangsu Bae, Sang-Yeon Lee

## Abstract

Mutations in nuclear genes that regulate the mitochondrial DNA (mtDNA) replication machinery have been linked to mtDNA depletion syndromes. Through whole-genome sequencing, we identified a heterozygous missense mutation (c.272G>A:p.Arg91Gln) in single-stranded binding protein 1 (SSBP1), a crucial protein involved in mtDNA replication. The proband manifested symptoms including sensorineural deafness, congenital cataract, optic atrophy, macular dystrophy, and myopathy, all of which are compatible with mtDNA depletion disease. We found that this *SSBP1* mutation impeded multimer formation and DNA-binding affinity, which led to reduced efficiency of mtDNA replication and altered mitochondria dynamics. To correct this mutation, we tested two adenine base editor (ABE) variants using patient-derived fibroblasts. One variant, NG-Cas9-based ABE8e (NG-ABE8e), exhibited higher editing efficacy (up to 30% editing), and edited cells showed signs of improved mitochondrial replication and function, including increased mtDNA and ATP production. However, these cells also had higher frequencies of off-target editing of nearby nucleotides, but the risks from bystander editing were limited due to mostly silent mutations and off-target sites in non-translated regions. The other variant, NG-Cas9-based ABE8eWQ (NG-ABE8eWQ), had a safer therapeutic profile with very few off-target effects, but this came at the cost of lower editing efficacy (up to 10% editing). Despite this, NG-ABE8eWQ-edited cells still restored replication and improved mtDNA copy number, which in turn recovery of compromised mitochondrial function. As these therapeutic strategies make their way into the clinic, our research suggests that base editing-based gene therapies may be a promising treatment for mitochondrial diseases, including those associated with *SSBP1* mutations.

## Introduction

Mitochondrial diseases are characterized by respiratory chain dysfunction and often manifest as multisystemic involvement, leading to a spectrum of symptoms^1^. These conditions often result from mutations in either the mitochondrial DNA (mtDNA) or nuclear genomes^2^. The mtDNA is a compact, double-stranded circular DNA with a high copy number. It encodes 37 essential genes, which include 13 core protein components of the mitochondrial respiratory chain complexes (Complexes I through V) as well as 2 ribosomal RNAs and 22 transfer RNAs necessary for their translation. Each cell contains thousands of copies of mtDNA, and these are organized into nucleoids, each carrying approximately a thousand molecules of the packaging protein and the replisome machinery^3^. Notably, the nuclear genome encodes over 99% of the mitochondrial proteome, encompassing not only respiratory chain subunits but also all the proteins responsible for mtDNA maintenance, replication, transcription and copy number control of the mitochondrial genome^4–8^. Mutations in these nuclear genes can induce instability of the mitochondrial genome, as characterized by mtDNA depletion and somatic accumulation of multiple deletions in postmitotic tissues, which in turn lead to mitochondrial diseases inherited in a Mendelian fashion^9^. While our understanding of mitochondrial biology and pathology has significantly advanced, the complex nature of mitochondrial inheritance combined with the contributions of nuclear and mitochondrial genomes has made it challenging to fully elucidate the mechanisms underlying mitochondrial diseases^10^.

Human mtDNA is replicated by specialized machinery distinct from the nuclear replisome^11^. In particular, the mitochondrial replisome, a complex multi-component apparatus, is pivotal for mtDNA replication^12^. Mutations in mitochondrial replication machinery can lead to mtDNA depletion, which can sometimes lead to deletions within the mitochondria genome^13^. One of components of the mtDNA replication complex, mitochondrial single-stranded DNA-binding protein 1 (SSBP1), is crucial for regulating mtDNA replication initiation and maintenance in mammalian mitochondria^14^. In humans, pathogenic mutations in *SSBP1* have been found to impair mtDNA replication fidelity and cause mtDNA depletion and/or deletion in mitochondria genome, which subsequently led to compromised oxidative phosphorylation and a spectrum of phenotypes in patients^15,16^. Until now, fewer than 15 mutations in *SSBP1* have been identified^17^ and the majority of these patients present with optic atrophy, sensorineural deafness, mitochondrial myopathy, and kidney failure.

Despite advancements in our understanding of the diagnosis and pathology of mitochondrial diseases, a definitive treatment remains elusive. Gene therapy has produced clinical benefits for several human diseases, and gene-editing technologies are expected to play a major role in the field’s future^18^. In particular, CRISPR-based technologies are currently being developed for mammalian genome editing to treat a number of genetic disorders^19^. The canonical CRISPR nucleases generate DNA double-stranded breaks (DSBs) at target sites, which are then repaired mostly by non-homologous end joining (NHEJ) and homology-directed repair (HDR). While HDR can correct disease-causing mutations, CRISPR-mediated DSBs can have unintended outcomes such as small insertions and/or deletions (indels) as well as large DNA deletions, chromosomal depletion, and p53-driven programmable cell death^20,21^. Alternatively, base editors that consist of Cas9 nickase and cytidine or adenosine deaminases were developed, which generate single-strand breaks instead of DSBs^20,22–25^. In general, cytosine base editors (CBE) and adenine base editors (ABE) can introduce C•G to T•A and A•T to G•C substitution, respectively, with high efficiency. Given that point mutations represent over half of all identified pathogenic genetic variants in humans^26^, base editors possess significant therapeutic potential for correcting disease-causing mutations. At present, base editors are undergoing clinical trials for several rare diseases^27^; thus mutations associated with mitochondrial diseases may be promising candidates for base editing-based gene therapy.

In this study, we identified a novel heterozygous mutation in *SSBP1* (c.272G>A:p.Arg91Gln) in one family exhibiting symptoms of sensorineural deafness, optic atrophy, macular dystrophy, early cataract, and mitochondrial myopathy, which are all compatible with mitochondria disease. We revealed that this *SSBP1* mutation altered multimer formation and reduced DNA-binding affinity, which compromised mtDNA replication fidelity and altered mitochondria dynamics. Ultimately, this *SSBP1* mutation led to mtDNA depletion and subsequent mitochondrial dysfunction. Our results replicate findings observed in *SSBP1* mutations. To rescue mitochondria function, we tested two ABE variants in patient cells harboring the *SSBP1* mutation and assessed editing efficacies, off-target effects, and functional recovery. Taken together, our findings suggest base editing-based gene therapies may be beneficial for mitochondrial diseases provide guidance for selecting appropriate base editors for potential clinical use.

## Materials and Methods

### Study subjects

We included the participants within the Hereditary Hearing Loss Clinic of the Center for Rare Diseases, Seoul National University Hospital, South Korea. All procedures were approved by the Institutional Review Board of Seoul National University Hospital (no. IRB-H-0905-041-281 and IRB-H-2202-045-1298). The demographic data and clinical phenotypes were retrieved from the electronic medical records.

### Molecular genetic testing

Genomic DNA was isolated from peripheral blood samples using the Chemagic 360 instrument (Perkin Elmer, Baesweiler, Germany). The genetic testing approach was organized into a sequential stepwise approach^28–32^. The details of molecular genetic testing were described in Supplementary Methods.

### Molecular modeling and structure analysis

The crystal structure of the wild-type SSBP1 protein was obtained from the Protein Data Bank (PDB ID: 6RUP). The mutagenesis was generated using PyMOL software (ver. 2.5.2). Additionally, the model structure of the SSBP1-ssDNA complex was generated, and the surface interface between SSBP1 and ssDNA was visualized by aligning the E-coli ssDNA structure (1EYG) to the SSBP1 crystal structure. Inter- and intrachain changes, including hydrophobic, hydrogen bonds, and salt bridge interactions, were compared to predict the structural effects of *SSBP1* mutation. All graphical illustrations were produced using PyMOL software (ver. 2.5.2) (PyMOL Molecular Graphics System ver. 2.0, Schrödinger Inc., New York, NY, USA).

### SDS-PAGE and immunoblotting

Cell lysates were mixed with sample buffer and denatured for SDS-PAGE. Proteins were then transferred to PVDF membranes, blocked, and incubated with primary and secondary antibodies. Protein bands were visualized using chemiluminescence. The comprehensive procedures are provided in the Supplementary Methods.

### Electrophoretic mobility shift assay

For the Electrophoretic Mobility Shift Assay (EMSA) targeting SSBP1, a specific ssDNA probe was employed. Both wild-type and mutant variants of SSBP1 proteins were combined with the probes and incubated for 30 minutes. Following this incubation, samples were subjected to electrophoresis on 2% agarose gels at 100 V for 45 minutes. Following electrophoresis, the gels were visualized using UV light. The detailed methods can be referenced in the Supplementary Methods.

### Mammalian cell cultures and transient expression

A549 cells were grown in RPMI 1640 supplemented with 10% of FBS, 100 U/mL penicillin, 100 μg/ml streptomycin, and 2 mM L-glutamine. A human *SSBP1* cDNA clone (RC215106) was purchased from Origene. The SSBP1 p.Arg91Gln mutation plasmid was generated utilizing the QuickChange mutagenesis method^33^. For transient overexpression, cells were transfected with 2 mg of total plasmid DNA in 6-well culture plate (>70-80% confluent of cells) for 24 h using Lipofectamine 3000 (L3000001, Invitrogen), according to manufacturer’s guidelines.

### Genomic DNA and RNA isolation followed by quantitative real-time PCR

Genomic DNA was purified from either A549 or fibroblasts using Puregene™ DNA purification kit (158026, QIAGEN). Concurrently, total RNA was isolated cells using TRIzol Reagent (15596018, Thermofisher). cDNA samples were prepared using one microgram of total RNA by reverse-transcription PCR (Accupower RT-pre-mix, Bioneer). qRT-PCR reactions were carried out using either the genomic DNA or cDNA, SYBR qPCR master mixture (RT501M, Enzynomics) as the reporter dye, and 10 pmol of primers to detect mRNA expression of specific genes. Primer sequences used in this study are summarized in Supplementary Table 3.

### Immunocytochemistry and EdU labeling assay

A549 cells were prepared for immunofluorescence microscopy, involving transfection, fixation, permeabilization, and antibody incubation. The cells were then mounted with DAPI-containing medium and imaged using a Leica STELLARIS 8 microscope. During EdU labeling, cells were treated with EdU, visualized with Alexa Fluor 488, and had their mitochondria stained with MitoTracker Red. Quantitative analysis involved counting EdU foci in images, normalized to cell size. Comprehensive methods can be found in the Supplementary Methods.

### Oxygen consumption rate

The Seahorse XF96 Extracellular Flux Analyser was employed to measure cellular oxygen consumption rate (OCR) and extracellular acidification rate (ECAR) in real time. Fibroblasts or A549 cells were seeded in Seahorse microplates and prepped with specific assay running media. Upon calibration, the analyzer measured OCR and ECAR simultaneously. The assay integrated four compound injections, including oligomycin, FCCP, rotenone, and antimycin A, to evaluate various mitochondrial functions like basal respiration, maximal respiration, and ATP production. The detailed procedures can be found in the Supplementary Methods.

### Fibroblast cell culture

A skin biopsy was taken from a donor under local anesthesia and stored in Phosphate Buffered Saline (PBS). This biopsy was subsequently sectioned into 9-12 distinct segments, which were then seeded in a 12-well plate supplemented with DMEM and 20% FBS. Upon reaching confluence, the fibroblasts were harvested for expansion. Detailed methodologies are provided in the Supplementary Methods.

### Electroporation and cell lysis

Patient-derived fibroblasts (100,000 cells/transfection) were transfected using a Neon transfection system 10μl kit (ThermoFisher, MPK1025) with following parameters: Voltage, 1600V; width, 20ms; number, 1. The fibroblasts were transfected with 900 ng of ABE-encoding plasmid and 300ng of sgRNA-encoding plasmid. After 72 hrs from electroporation, half of the cells were harvested for next-generation sequencing (NGS) and the other half of the cells were maintained for verifying functional recovery. Cell pellets were resuspended in proteinase K extraction buffer containing 40mM Tris-HCl [pH8.0], 1% Tween 20, 0.2mM EDTA, 10mg of Proteinase K, 0.2% Nonidet P-40 (VWR Life Science, 97064-730). After resuspension, incubation was performed at 60°C for 15min and 98°C for 5min.

### High-throughput DNA sequencing

PCR amplification was carried out with primers containing sequencing adaptors using KOD-Multi & Epi (TOYOBO, KME-101) according to the manufacturer’s instructions. After the first PCR amplification, 1μl of the 1st PCR products were amplified again with primers containing sequencing barcodes of TruSeq HT Dual index system (Illumina). Then, the 2nd PCR products were purified using Expin PCR SV mini (GeneAll) and sequenced using a Miniseq sequencing system (Illumina). The sequencing results were analyzed using BE-Analyzer^34^. Analysis of sequencing data was mostly carried out using the computing server at the Genomic Medicine Institute Research Service Center.

### Statistical analysis

We conducted statistical analysis and generated graphs using GraphPad Prism 8.0.1 (GraphPad Software). Immunoblots and gel images were quantified using ImageJ software. Specific experiment details can be found in the figure legends. All experimental data is presented as mean ± standard error of the mean (SEM). Statistical significance was determined for comparisons with a P value < 0.05, denoted as *, *p* < 0.05; **, *p* < 0.01; and ***, *p* < 0.005.

## Results

### Identification of *SSBP1* mutation and associated clinical phenotype

Patient under 5 years of age was a sporadic case in the SNUH family, exhibiting symmetrical moderate-to-severe, down-sloping sensorineural deafness. This was confirmed via electrophysiological testing, including auditory brainstem response threshold and auditory steady-state response (**Figures 1a and 1b**). Both transient-evoked otoacoustic emissions (TEOAEs) and distortion product otoacoustic emissions (DPOAEs) were undetectable across all frequencies (**Figure 1c**) and the sensorineural deafness progressively deteriorated over the next two years. Neither temporal bone computed tomography nor brain magnetic resonance imaging revealed any inner ear or brain anomalies (data not shown). Additionally, tests for congenital cytomegalovirus (cCMV) infection yielded normal results. Using a stepwise genomic approach, we performed trio whole-genome sequencing (WGS) and identified a *de novo* heterozygous missense mutation (c.272G>A:p.Arg91Gln) in the *SSBP1* gene. Sanger sequencing in parental samples confirmed that the mutation arose *de novo* (**Figure 1d**). We examined several population databases, including Genome Aggregation Database (gnomAD) and the Korean Variant Archive (KOVA), but could not find any reports of this missense mutation (p.Arg91Gln), which was located in the single-stranded binding (SSB) domain. The Arg91 residue is conserved across *SSBP1* orthologs of multiple species (http://genome.ucsc.edu/) (**Figure 1e**). In silico analyses also predicted this mutation was disease-causing, with a Combined Annotation Dependent Depletion (CADD) phred score of 23.9. Based on the American College of Medical Genetics and Genomics/Association for Molecular Pathology (ACMG/AMP) guidelines^35^, the *SSBP1* mutation (c.272G>A:p.Arg91Gln) is also considered “pathogenic” (**Supplementary Table 1**).

**Figure 1.**
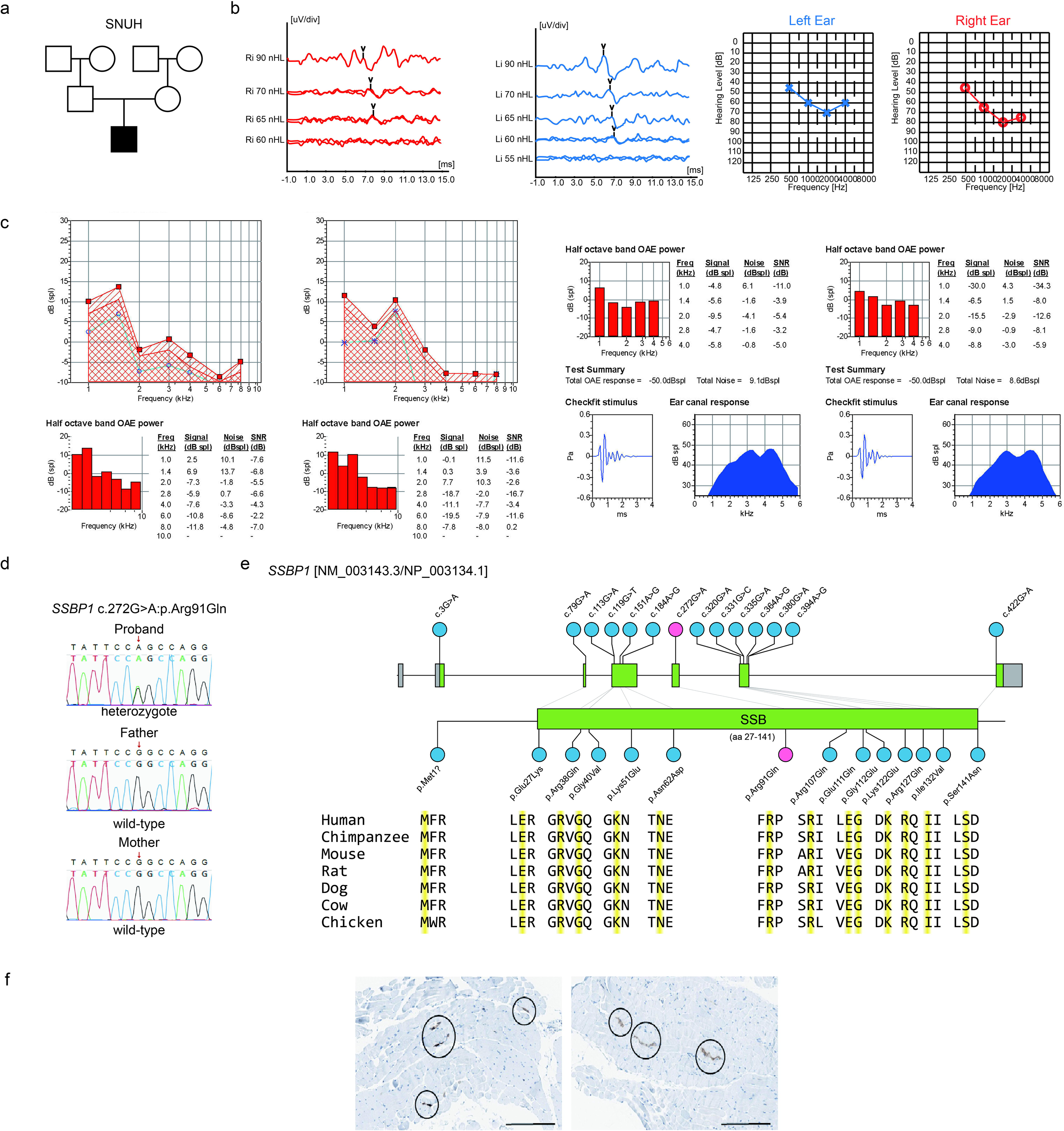
Characterization of *de novo* mutation in *SSBP1*. (a) The pedigree of the Korean SNUH family harboring a *de novo* heterozygous *SSBP1* mutation (c.272G>A:p.Arg91Gln). Filled symbols and opened symbols indicate affected and unaffected individuals, respectively. (b) auditory brainstem response threshold (ABRT, left) and auditory steady-state response (ASSR, right) reveal a bilateral symmetric, moderately severe sensorineural deafness. (c) Neither transient evoked otoacoustic emission (TEOAE) nor Distortion product otoacoustic emission (DPOAE) showed responses across all tested frequencies, suggesting its loss of outer hair cell in the cochlea. (d) Sanger sequencing chromatograms of the *SSBP1* c.272G>A:p.Arg91Gln mutation in the SNUH family. The arrow indicates the site of the mutation. (e) The sequence domain (upper) and conservation maps (lower) showcase *SSBP1* mutations previously reported in the literature (blue circles), with the inclusion of the novel mutation (c.272G>A:p.Arg91Gln) from this study (red circle). In the conservation map, gray regions denote untranslated regions, while green regions signify the coding sequence. All the mutations’ residues, including Arg91 residue, were highly conserved among the *SSBP1* orthologs in various species (highlighted in yellow). (f) Immunohistology images of CD56 staining of left thigh muscle tissue taken from proband’s muscle biopsy. Ultrathin sections revealed myofibers with mild size variation, including both degenerating and regenerating myofibers, which predominantly appear rounded. Within the subsarcolemmal area, a few myelin, glycogen particles, and fat vacuoles are discernible. A moderate degree of endomysial fibrosis is evident. The presence of CD56-positive cells (black circles), indicating degenerating myofibers (16/10 HPF), was also noted. These findings suggest a mild myopathic change, consistent with mitochondrial myopathy.

In the literature, ten studies have identified thirteen distinct missense mutations in *SSBP1* (**Figure 1e** **and Supplementary Table 2**), all of which are associated with mtDNA depletion syndrome. Although optic atrophy is the predominant phenotype observed in patients with *SSBP1* mutations, a variety of additional neurological symptoms are commonly reported, including sensorineural deafness (**Supplementary** Figure 1). In mouse cochlea (P5), we observed that *SSBP1* is ubiquitously expressed, including the hair cells of the organ of Corti, stria vascularis, and spiral ganglion neurons, which are essential for sound transduction (**Supplementary** Figure 2). Genetic diagnosis can guide a complete workup by providing additional clinical options for the patient such as targeted agents, referral to specialists, tailored auditory rehabilitation, or symptomatic treatment^36^. Given this, our patient was referred to subspecialists, including a pediatric ophthalmologist to evaluate optic neuropathy with retinal abnormalities, and pediatric neurologists to evaluate neurodevelopmental delay and other associated muscular symptoms. The initial outpatient examination revealed no abnormal findings in the cornea or conjunctiva. However, upon slit-lamp examination and ultrasonography, remarkable lens opacity affecting the central visual axis was observed in both eyes (**data not shown**). Visual acuity loss was marked only with the fixation of large objects in both eyes. In the fundus examination, lens opacity resulted in a blurry appearance of the macula, which is an important anatomical area for central vision (**data not shown**). The patient underwent bilateral lensectomy and anterior vitrectomy at one-month intervals. At the four to five months post-operative follow-up examination, the Snellen visual acuity improved to 20/32 in the right eye and 20/40 in the left eye. Slit lamp examination and ultrasonography confirmed clear removal of lens opacity (**data not shown**). No other post-surgery complications, such as retinal traction or vitreous hemorrhage, were observed, apart from mild vitreous opacity. Fundus examination revealed optic disc atrophy and degeneration of the retinal pigment epithelium in the macula (**data not shown**). Additional follow-up by neurologists identified mild muscle weakness, which was consistent with the detection of CD56-positive degenerated myocytes by muscle biopsy (**Figure 1f**). These results suggested a mild mitochondrial myopathy, although overt muscle phenotypes were not observed, and creatine kinase (CK) levels remained within the normal range. Overall, the patient exhibited signs of sensorineural deafness, congenital cataract, optic atrophy, macular dystrophy, and myopathy, all of which were indicative of mitochondrial disease.

In the literature review, mtDNA depletion has been observed in the majority of patients with mitochondrial disease (**Supplementary Table 2**), whereas mtDNA deletions have been specifically reported in cases with double hits (*SSBP1* c.3G>A and *MT-RNR1* m.1555A>G)^37^ and in a patient with a *SSBP1* c.79G>A mutation^38^. In our patient, neither mtDNA panel sequencing nor Multiplex genomic Ligation-dependent Probe Amplification (MLPA) of genomic DNA extracted from peripheral blood samples and patient-derived fibroblasts identified any pathogenic mtDNA variants or copy number variations (CNVs) in mitochondria genome (**Supplementary Data 1 and Supplementary** Figure 3). Additionally, long-range PCR performed on skeletal muscle confirmed the absence of mtDNA deletions (**Supplementary** Figure 4).

### Overexpression of mutant SSBP1 decreases mtDNA copy number

To explore the molecular consequences of the *SSBP1* c.272G>A mutation, we first assessed the impact of overexpressing Flag-tagged wild-type or p.Arg91Gln mutant protein on mtDNA copy number. By performing quantitative PCR analysis on mtDNA-specific genes *ND1* and *ND5* and normalizing it to *SLCO2B1* or *SERPINA1*^39^, we observed a significant decrease in mtDNA copy number in transiently overexpressed mutants compared to the wild-type protein (**Figure 2a**). However, in mitochondria isolated from A549 cells, SSBP1 protein was similarly expressed in the p.Arg91Gln mutant and the wild-type (**Figure 2b**). Subsequent quantitative real time PCR (qRT-PCR) also indicated there was no significant difference in *SSBP1* mRNA between transient overexpression of SSBP1 wild-type and the p.Arg91Gln mutant (**Figure 2c**). We also performed an immunofluorescence assay to visualize the colocalization of SSBP1 with MitoTracker Red-labeled mitochondria. We found a significant overlap between SSBP1-positive Flag signals and mitochondrial networks, confirming SSBP1 is a mitochondrial protein (**Supplementary** Figure 5). Our findings suggest that the transcription and translation of the SSBP1 p.Arg91Gln mutant is similar to its wild-type counterpart. We therefore hypothesized that the subsequent decrease in mtDNA copy number may be due to a functional deficiency.

**Figure 2.**
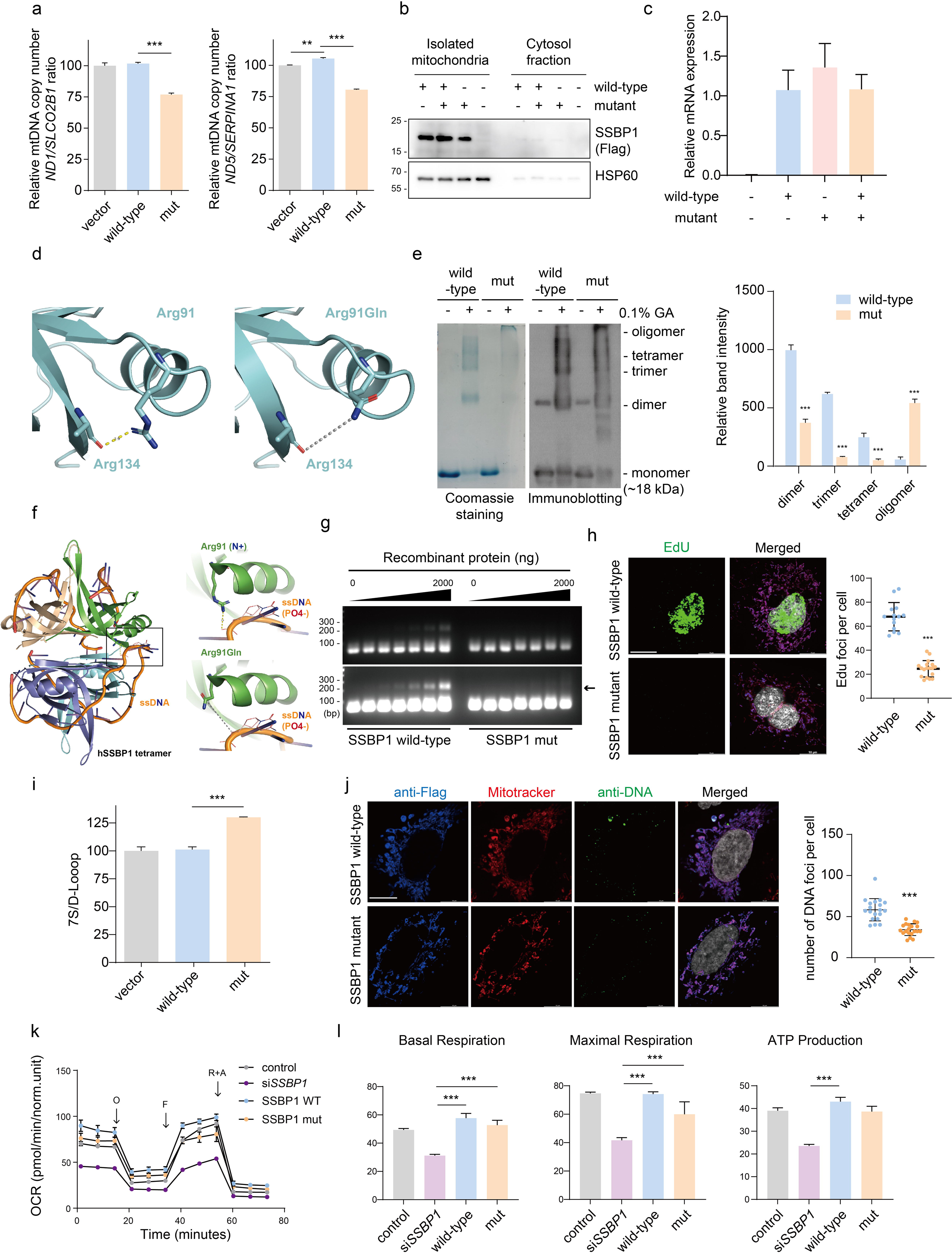
Overexpression of mutant *SSBP1* induces mtDNA depletion and mitochondrial dysfunction in A549 cells through the inhibition of mtDNA replication. (a) Quantification of mtDNA was performed using genomic DNA isolated from A549 cells overexpressing *SSBP1*. Relative copy numbers of mtDNA genes *ND1* and *ND5* were measured using qRT-PCR. *SLCO2B1* and *SERP1NA* DNA levels were used to normalize the results (mean ± SEM, n = 3 independent experiments). (b) Immunoblots for Flag-tagged *SSBP1* and HSP60 in mitochondrial and cytosolic fractions isolated from A549 cells overexpressing wild-type and mutant *SSBP1*. (c) qRT-PCR of *SSBP1* mRNA expression in A549 cells overexpressing wild-type and mutant *SSBP1*. *GAPDH* mRNA was used to normalize the results (mean ± SEM, n = 3 independent experiments). (d) Ribbon diagram showing intrachain interaction of *SSBP1* Arg91 with Ala134 (PDB 3ULL). The Ala134 residue forms a hydrogen bond with the Arg91 residue; however, the interaction with the Ala134 residue was lost with the Arg91Gln mutant. (e). Images of Coomassie Brilliant Blue (CBB) staining (left) and *SSBP1* immunoblotting for *SSBP1* recombinant proteins after oligomerization. Each immunoblot band intensity was quantified using ImageJ (means ± SEM, n = 3 independent experiments, unpaired Student’s *t* test). (f) Ribbon diagram of *SSBP1* tetramer structure with aligned ssDNA (PDB 1EYG). The negatively charged ssDNA interacts with the positively charged Arg91 residue; however, the Arg91Gln residue is predicted not to interact. (g) Electrophoresis Mobility Shift Assay (EMSA) of *SSBP1*. ssDNA probe (10 μM) was incubated with increasing amounts (0, 0.07, 0.13, 0.25, 0.5, 1, and 2 μg) of wild-type and mutant recombinant *SSBP1* protein. The arrow denotes the shifted form. (h) Representative confocal images of A549 cells with *SSBP1* overexpression with EdU incorporation (green). *SSBP1* was stained using Anti-Flag (blue), while mitochondria were stained with MitoTracker (red) (also see Supplementary Figure 7) and nuclei were stained with DAPI (white). The number of EdU foci per cell was counted manually (means ± SEM, n = 14-18, unpaired Student’s *t* test). Scale bar = 10 μm. (i) 7S DNA was quantified in A549 cells overexpressing *SSBP1* using qRT-PCR. The results were normalized to mtDNA levels (means ± SEM, n = 3 independent experiments, unpaired Student’s *t* test). (j) Immunofluorescence images of Flag-tagged *SSBP1* (anti-Flag, blue) in A549 cells overexpressing wild-type or mutant *SSBP1*. Mitochondria were detected using MitoTracker (red), and mtDNA was stained with anti-DNA (green). The number of nucleoid per cell was counted manually (means ± SEM, n = 19-20, unpaired Student’s *t* test). Scale bar = 10 μm. (k) Oxygen consumption rate (OCR) in A549 cells overexpressing *SSBP1* wild-type and mutant under basal conditions and after injection of oligomycin (O), carbonylcyanide 4-(trifluoromethoxy) phenylhydrazone (FCCP; F), rotenone (R) and antimycin A (AA). (l) Basal respiration, maximal respiration, and ATP production in A549 cells were calculated from OCR traces and are reported in the graph (mean ± SEM, n = 5, unpaired Student’s *t* test).

### *SSBP1* c.272G>A mutation impedes multimer formation and disrupts DNA-binding affinity

In humans, wild-type SSBP1 protein forms a stable tetramer formed from two dimers^17^. Using the crystal structure of SSBP1 (PDB 3ULL), we examined the impact of the p.Arg91Gln mutation on protein stability and multimerization. While the Arg91 residue is not involved in interchain interactions, the substitution to Gln91 disrupts the hydrogen bond between the NH+ of Arg91 and the C=O of Ala134 of the same protomer (**Figure 2d**). To examine the effect of the mutation in SSBP1 oligomerization, we performed glutaraldehyde (GA) crosslinking experiments on wild-type and mutant proteins (**Figure 2e**). In contrast to the wild-type protein, which exhibited typical multimer formation including di-, tri-, and tetramers, the mutant showed a reduced capacity to form each of these multimers, with only 37.4%, 12.7%, and 20.5% of dimers, trimers, and tetramers formed, respectively, as compared to wild-type. In addition, the mutant protein showed more high oligomeric species indicating that the mutant protein might form aggregates due to its lower stability.

Next, we tested the binding strength of both wild-type and mutant proteins to ssDNA. During replication, SSBP1 directly binds to single-stranded mtDNA in its tetramer form to prevent the reannealing of strands and protect against nucleolytic attacks^15,40^. Through molecular modeling of the SSBP1-ssDNA complex, we speculated that our mutation would reduce DNA-binding affinity by removing the ionic interaction between the positively charged Arg91 and the negatively charged DNA phosphate backbone (**Figure 2f**), possibly leading to reduced DNA-binding affinity. Although SSBP1 has been shown to directly bind to DNA^41^, the molecular interaction between the protein and the DNA is not well understood due to a lack of structural information. We therefore proceeded to examine the DNA-binding affinity of the *SSBP1* mutation. Electromobility shift assay (EMSA) demonstrated that our ssDNA probe interacted specifically with wild-type SSBP1 but not the Arg91Gln mutant in a dose-dependent manner, indicating the Arg91Gln mutation significantly decreases DNA binding (**Figure 2g** and **Supplementary** Figure 6). In summary, the *SSBP1* mutation identified in our patient appears to impede multimer formation and disrupt DNA-binding affinity without affecting the abundance of SSBP1 itself. Considering both multimer formation and DNA-binding affinity play pivotal roles in ensuring proper mtDNA maintenance^42^, we hypothesize that this mutation compromise mtDNA replication efficiency, which has been associated with aberrant mtDNA maintenance.

### *SSBP1* c.272G>A mutation compromises mtDNA replication fidelity and alters mitochondrial network dynamics

To evaluate the effect of the *SSBP1* c.272G>A mutation on the efficiency of mtDNA replication, we performed an EdU incorporation assay to visualize and quantify mtDNA synthesis in our transient overexpression system. We found that in cells expressing the mutated SSBP1, the co-localization of EdU foci in mitochondria (i.e., foci within the mitochondria) occurred at 36.1% of the amount observed in wild-type cells, indicating that mtDNA replication was reduced. (**Figure 2h** and **Supplementary** Figure 7). Consistent with this result, we also observed a significant rise in 7S DNA levels in cells harboring the mutation (**Figure 2i**). Considering the role of 7S DNA in the replication machinery^43^, increased 7S DNA in mutant cells may reflect incomplete mtDNA replication and could be a compensatory response to the decreased replication rate.

The mtDNA replication machinery and mitochondrial maintenance are known to be interrelated^44^. We subsequently investigated the number of nucleoids colocalizing with mitochondria labeled by MitoTracker Red. Our data revealed a 42% reduction in anti-DNA immunofluorescence in the mutant cells relative to the wild-type cells (**Figure 2j**), which agreed with the observed decrease in mtDNA copy number in mutant SSBP1-transfected cells. Additionally, we visualized the dynamics of mitochondrial network using the MitoTracker Red. The A549 cells overexpressing the SSBP1 mutant displayed more pronounced fragmentation and a shortened morphology compared to wild-type cells (**Figure 2j**). These observations imply that the *SSBP1* mutation may cause changes in mitochondrial network dynamics due to compromised mtDNA replication and maintenance.

### mtDNA depletion influences bioenergetics in *SSBP1* mutant cells

To explore the ramifications of mtDNA depletion on cellular metabolism, we examined mitochondrial function using Seahorse assays. In our assessment of the OCR, we observed reduced respiratory function in cells overexpressing the SSBP1 mutant compared to those with wild-type SSBP1 (**Figure 2k**). The reduction in OCR for the mutant cells was consistently observed. Moreover, this decrease became more pronounced upon endogenous SSBP1 knockdown in wild-type cells by siRNA, with significant reductions in basal respiration, maximal respiration, and ATP production (**Figure 2l**). In addition, we confirmed that the expression level of oxidative phosphorylation system (OXPHOS) complex were reduced in cells overexpressing the SSBP1 mutant compared to wild-type SSBP1 (**Supplementary** Figure 8). These findings suggest that SSBP1 may contribute to mitochondrial ATP synthesis in addition to its role in mtDNA replication.

### Correction of *SSBP1* mutation using ABE variants to balance editing efficiency and off-target effects

To correct the heterozygous *SSBP1* point mutation (c.272G>A:p.Arg91Gln) in patient-derived fibroblasts, we used the Cas9-based editor ABE8e that targeted a “NG” protospacer adjacent motif (NG-ABE8e) with two different single-guide RNAs (NG-sg1, NG-sg2), or a canonical Cas9-based ABE8e with one sgRNA (NGG-sg1). While the NG-sg2 and NGG-sg1 demonstrated a very poor A-to-G substitution rate (average 53.46% and 53.4% respectively, including normal allele frequency), NG-sg1 showed the highest A-to-G substitution rate (average 75% including normal allele frequency) (**Figure 3a**). Therefore, we chose to focus on NG-sg1 to correct the missense mutation. Although NG-ABE8e with NG-sg1 corrected the target A with high frequency, it also showed some bystander editing. These edits included nucleotides near the target A (A5, by counting the end distal to the PAM as position 1), adenines (A-1, A9) and cytosines (C7, C8) at the following frequencies: A-1: 0.29 ± 0.27%, A9: 14.99 ± 3.86%, C7: 0.87 ± 0.44% and C8: 0.79 ± 0.63% (**Figure 3b**). Fortunately, the conversion of either bystander A (A-1, A9) to G leads to a silent mutation that does not alter the amino acid sequence of SSBP1. In contrast, bystander C editing (C7, C8) can generate additional missense mutations (Pro92Ser for C7 substitution, Pro92Leu for C8 or both C7, C8 substitution) due to off-target activities of ABEs^45^. Although the frequency of bystander C editing was low, we tested a different ABE variant with mutations at V106W and D108Q (NG-ABE8eWQ) that has been shown to minimize bystander C-to-G editing by using a narrower targeting window^46^. As expected, bystander C editing was reduced; however, this also compromised target A (A5) editing efficiency (average 57.09± 3.17%, including normal allele frequency) (**Figure 3b**).

**Figure 3.**
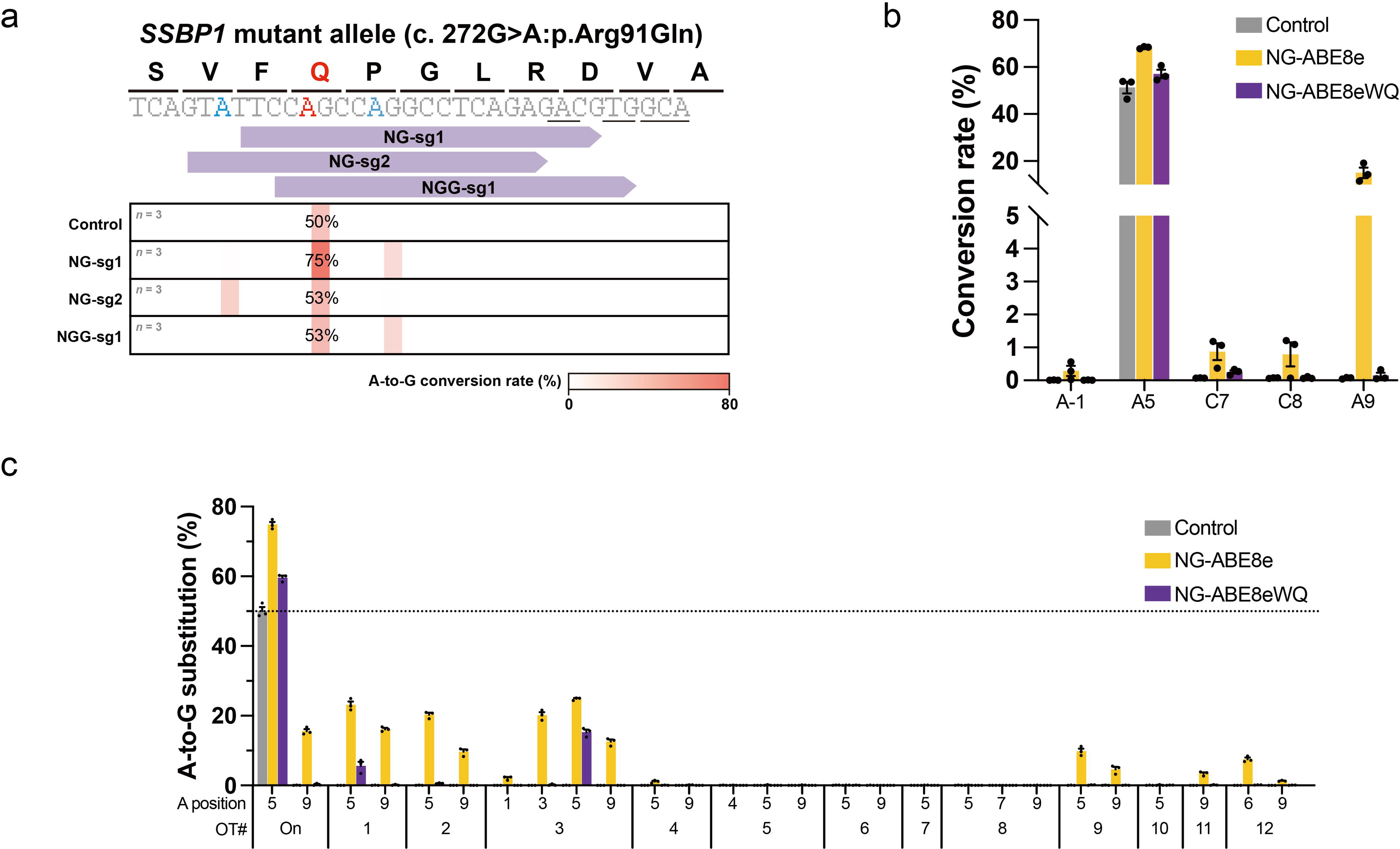
Analysis of sgRNA and ABE variant efficacy for *SSBP1* mutation and their off-target effects. (a) Positions and editing efficiencies of sgRNA candidates within the *SSBP1* gene. The red colored A indicates the target nucleotide (c.272G>A) and blue colored A indicates bystander As. A-to-G conversion rate for each nucleotide is shown using heatmaps. Underlined base identifies the PAM of each sgRNA. The editing frequency displayed on heatmap is an average value between three experimental replicates. (b) A-to-G or C-to-other conversion rate after using NG-sg1. The position of base is defined as counting the end distal to the PAM as position 1. (c) sgRNA-dependent off-target results investigated by Cas-OFFinder software. Dashed line indicates 50% frequency because of the heterozygosity of on-target.

We also investigated sgRNA-dependent off-target effects from NG-sg1. We used Cas-OFFinder software to identify 12 potential off-target sites (OT1-OT12) when up to three mismatched bases were allowed^47,48^. Because NG-ABE8e has a relatively wide editing window and high deaminase activity, NG-ABE8e showed substantial off-target editing frequency at OT1-OT3, OT9, OT11-OT12 (**Figure 3c**). OT1-OT3 and OT11 are in non-coding regions, whereas OT9 and OT12 are in intron regions within the genes *MPDZ* and *DOCK3*, respectively. Meanwhile, NG-ABE8eWQ showed significantly reduced off-target editing (**Figure 3c**). These results suggest that NG-ABE8e may present the highest functional recovery and the risks from bystander editing may be limited due to mostly silent mutations and off-target sites in non-translated regions. In contrast, our use of the safer ABE variant NG-ABE8eWQ also corrected the *SSBP1* mutation, but with reduced efficiency.

### ABEs improve mtDNA copy number and replication efficiency in patient-derived fibroblasts

We next evaluated the efficacy of these ABE variants (NG-ABE8e and NG-ABE8eWQ) to restore mtDNA copy number and mitochondrial replication. We performed genome editing in patient-derived fibroblasts and compared them to two control cell lines from healthy individuals without *SSBP1* mutations (control #1), and parental control cells (control #2). One concern with assessing mitochondrial function in cell culture can be the potential confounding effects from cellular aging and senescence^49,50^. Our observations showed that patient fibroblast cells displayed substantial cellular senescence (β-galactosidase staining and swelling) after passage 7, whereas control and genome-edited cells had a limited number of senescent cells (**Supplementary** Figure 9). We therefore used fibroblast cells between passages 4 and 7 in our genome editing experiments to limit these age-dependent effects.

First, we examined SSBP1 protein expression in our fibroblast cell lines. Consistent with the data from the overexpression system, the levels of SSBP1 protein appeared to be comparable between groups **(Supplementary** Figure 10). We then quantified the mtDNA copy number using qRT-PCR for mtDNA-encoded *ND1* and *ND5* and normalized it to *SLCO2B1* or *SERP1NA1*. The mtDNA copy number in patient fibroblast cells was significantly reduced compared to control fibroblasts, whereas the genome-edited cells consistently demonstrated a substantial recovery in mtDNA copy number (**Figure 4a**). On average, the mtDNA content in genome-edited cells increased by 59.9% with NG-ABE8e and 30.2% with NG-ABE8eWQ. In line with this, the relative number of nucleoids colocalizing with mitochondria was markedly reduced in patient fibroblasts but was significantly ameliorated in the genome-edited cells (**Figure 4b**). In the edited fibroblasts, the nucleoid ratio increased by 1.5-fold with NG-ABE8e and 1.3-fold with NG-ABE8eWQ compared to patient cells. These levels correspond to 94.0% and 79.1%, respectively, of those observed in control cells (**Figure 4b**).

**Figure 4.**
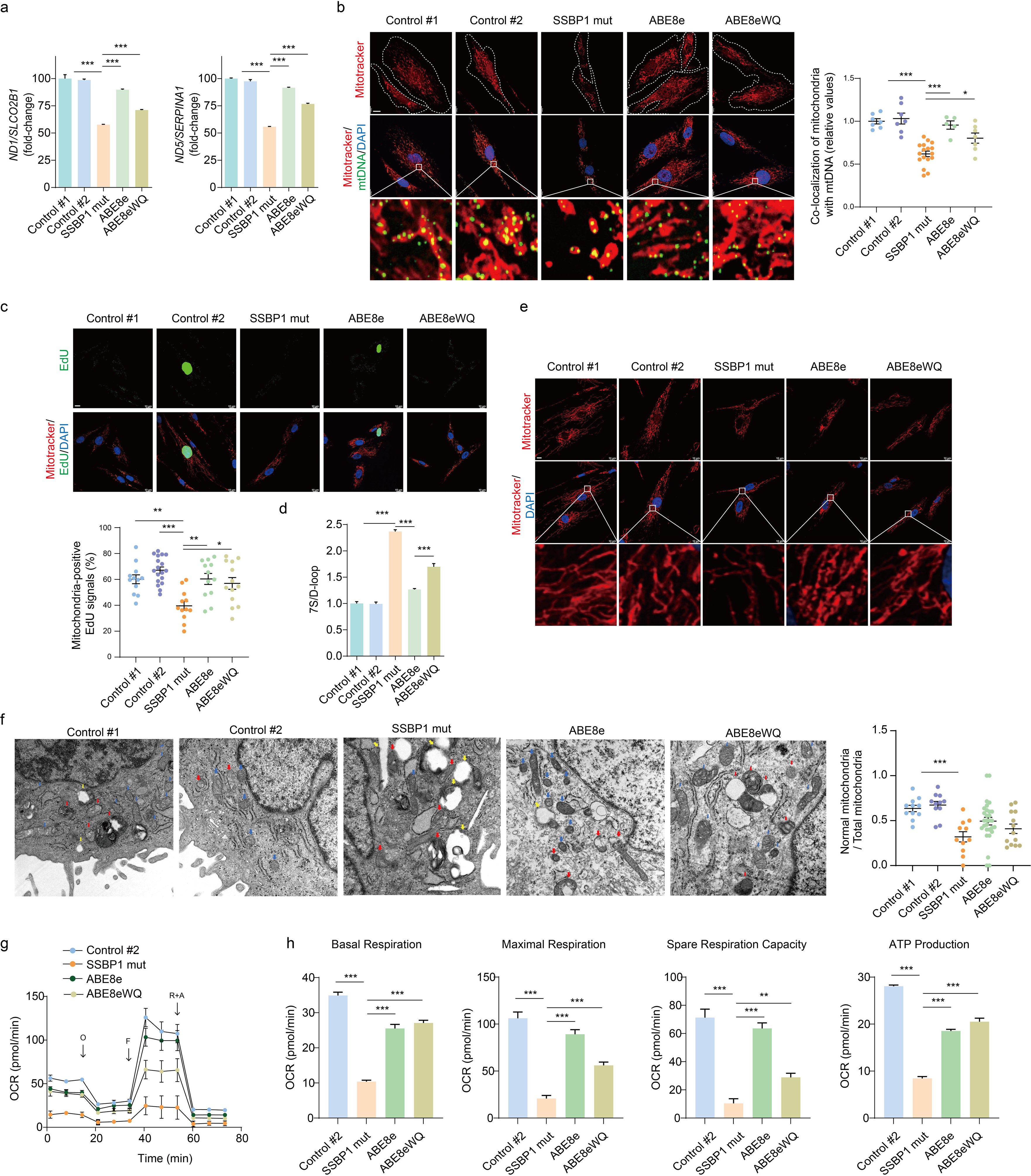
Functional recovery of mitochondria in patient-derived fibroblasts using ABEs. (a) Genomic DNA was isolated from the fibroblast cell lines and subjected to quantitative real-time PCR for *ND1* and *ND5*. *SLCO2B1* or *SERP1NA* was used to normalize the results (means ± SEM, n = 3 independent experiments). (b) Representative confocal images showed control, patient-derived, and genome-edited fibroblasts stained with MitoTracker (red) and anti-DNA (green). The boxes in the merged images point to the enlarged sections displayed at the bottom of each panel. The relative amounts of DNA that co-localized with MitoTracker were quantified (means ± SEM, n = 5-17, one-way ANOVA followed by the Bonferroni *post hoc* test). Scale bar = 10 μm. (c) Same as (b), except that immunofluorescence with with EdU incorporation. The co-localization of MitoTracker (red) and EdU (green) foci was employed to confirm the replication of mtDNA. The EdU incorporation into the mitochondria of different fibroblast cell lines was calculated and statistically analyzed (means ± SEM, n = 12-18, one-way ANOVA followed by the Bonferroni *post hoc* test). Scale bar = 10 μm. (d) Quantification of 7S DNA within the genomic DNA from fibroblast cell lines by qRT-PCR. The mtDNA level was used to normalize the result (means ± SEM, n = 3 independent experiments, one-way ANOVA followed by the Bonferroni *post hoc* test). (e) Representative confocal images depict mitochondrial fission in fibroblast cell lines. Mitochondrial network formation was assessed through MitoTracker staining (red). The boxes in the merged images indicate the enlarged sections shown at the bottom of each panel. Scale bar = 10 μm. (f) Transmission electron microscopy images of mitochondria with normal (blue arrow), large vacuole (yellow arrow), and abnormal (red) morphology in fibroblast cell lines. The relative proportion of normal mitochondria within the total mitochondria population was calculated (means ± SEM, n = 7-29, one-way ANOVA followed by the Bonferroni *post hoc* test). (g) OCR in fibroblasts under basal conditions and after injection of oligomycin (O), carbonylcyanide 4-(trifluoromethoxy) phenylhydrazone (FCCP; F), rotenone (R) and antimycin A (AA). (h) Alterations in basal respiration, maximal respiration, and ATP production in fibroblasts were derived from OCR traces (mean ± SEM, n = 3-6 one-way ANOVA followed by the Bonferroni *post hoc* test).

We subsequently evaluated if ABEs enhance mtDNA replication efficiency in patient fibroblasts by visualizing the EdU signal colocalized with mitochondria, which would suggest restoration of mtDNA copy number. Patient fibroblasts exhibited a significant reduction in the number of EdU foci that colocalized with mitochondria (**Figure 4c**). In contrast, genome-edited cells with NG-ABE8e and NG-ABE8eWQ demonstrated an increase in these EdU foci by 94.8% and 89.4%, respectively, compared to control fibroblasts (**Figures 4c**). Consistent with this observation, the elevated 7S DNA in edited patient fibroblasts was reduced by 47.0% or 28.0% when using NG-ABE8e or NG-ABE8eWQ, respectively, relative to patient fibroblasts (**Figures 4d**). The improvement in mtDNA replication fidelity through base editing, coupled with compensatory adjustments in 7S DNA levels, suggests that mutant SSBP1 may have a causal role in mtDNA depletion disease. These results imply that base editors-based gene therapies may be able to restore replication and improve mtDNA copy number, which in turn may contribute to the recovery of compromised mitochondrial function and abnormal dynamics observed in patient cells.

### ABE variants improve bioenergetics and mitochondrial dynamics

Next, we compared the mtDNA integrity among control groups, our patient-derived fibroblast, and ABE8e-modified patient-derived fibroblast. Similar to our overexpression system, mtDNA depletion also compromised the mitochondrial network in our patient-derived fibroblasts, as characterized by pronounced fragmentation and mitochondrial shortening (**Figure 4e**). Interestingly, NG-ABE8e- or NG-ABE8eWQ-treated patient-derived fibroblasts exhibited significant improvements in these dynamics (**Figure 4e**). To further elucidate the impact of ABEs on mitochondrial morphology and abundance, we examined the ultrastructural features of mitochondria by transmission electron microscopy (TEM) (**Figure 4f**). Alterations in mitochondrial fusion and fission have been associated with changes in mitochondrial shape and number, particularly inner membrane (i.e., cristae) remodeling during apoptosis^51^. In this study, abnormal mitochondrial morphology was defined as either vesicular or swollen forms. Normal mitochondria are characterized by dense staining of the cristae alongside an intact outer membrane. In contrast, vesicular mitochondria display morphological features where the cristae are separated from the inner membrane, with circular or rounded cristae distributed throughout the mitochondrial body. The swollen mitochondria exhibit fragmented or disorganized cristae, with large vacuole features^51^. In patient fibroblasts, the overall mitochondrial ultrastructure was abnormal, showing vesicular, swollen, or vesicular-swollen forms (**Figure 4f**). Furthermore, the patient-derived cells exhibited a substantial reduction in the proportion of normal mitochondria, corresponding to 48.9% of the control fibroblast levels. In contrast, the edited cells displayed a notable increase in the proportion of normal mitochondria, which corresponded to 75.5% and 62.6% of control fibroblast levels for NG-ABE8e and ABE8eWQ, respectively, with no statistically significant difference between the two base editors (**Figure 4f**).

We next investigated the impact of base editors on SSBP1-dependent bioenergetics. The OCR assay showed a markedly reduced respiratory capacity in patient fibroblasts compared to controls (**Figure 4g**). In contrast, treatment with either of the two ABEs significantly enhanced basal respiration (p<0.001 to NG-ABE8e; p<0.001 to NG-ABE8eWQ), maximal respiration (p<0.001 to NG-ABE8e; p<0.001 to NG-ABE8eWQ), spare respiration capacity (p<0.001 to NG-ABE8e; p=0.029 to NG-ABE8eWQ) and mitochondrial ATP production (p<0.001 to ABE8e; p<0.001 to ABE8eWQ), compared to patient cells (**Figures 4h**). In the edited cells, both ABEs exhibited enhanced basal respiration, with values of 2.46-fold for NG-ABE8e and 2.61-fold for NG-ABE8eWQ when compared to patient cells. This corresponds to 73.0% and 77.6% of the control cell levels, respectively. These changes in respiratory capacity led to increased ATP synthesis by OXPHOS, with increases of 2.01-fold for NG-ABE8e and 2.11-fold for NG-ABE8eWQ when compared to patient cells (**Figure 4h**). These values represent 66.1% and 73.1% of the control cell levels, respectively. Additionally, the spare respiratory capacity in the edited cells showed a marked increase (3.12-fold for NG-ABE8e and 2.08-fold for NG-ABE8eWQ) in comparison to patient cells (**Figure 4h**). These increases correspond to 89.2% and 40.5% of control cell levels, respectively. Such an enhanced spare respiratory capacity in the edited cells suggests that their mitochondria may have the ability to respond to increased energy demands or metabolic stress. In summary, these findings suggest that even when a base editor was less efficient at correcting a mutation (here *SSBP1*), it could still substantially improve mitochondrial function, including bioenergetics and mitochondrial dynamics.

### *SSBP1* disease-causing mutations are suitable for base editing

In our literature review (**Supplementary Table 2**), we noted that all of the documented *SSBP1* mutations with mitochondrial phenotypes were single nucleotide variations (SNVs). Therefore, base editors could potentially target all of these mutations. 89.5% and 8.7% of all patients with *SSBP1* mutations have either G-to-A or A-to-G single base transition mutations, which can be corrected by ABE and CBE, respectively. The remaining 1.8% of patients have either G-to-T or G-to-C single base transversion mutations (**Figure 5a**), which could potentially be corrected by more recently developed base editors. The A-to-Y base editor (AYBE), described by the Yang group, can induce A-to-C or A-to-T substitution to edit G-to-T mutations^52^. For G-to-C mutations, the C-to-G base editor (CGBE), characterized by the Joung group, can be used^53^. Although all *SSBP1* disease-causing mutations can theoretically be corrected using a variety of base editors, the majority of pathogenic *SSBP1* mutations are G-to-A transitions that can be corrected by ABE. We therefore used the DeepBE deep learning model, which can predict base editing efficacy in silico^54^, to simulate the potentially efficacy of ABEs to correct this type of *SSBP1* mutation. The prediction was performed based on the ABE8eW with diverse Cas9 variants. The results from this model showed all other mutations examined a more favorable base editing rescue score in comparison to c.272G>A, which was the original mutation we identified in this study (**Figure 5b**). Given the substantial improvement in mitochondrial function we observed after correcting the *SSBP1* c.272G>A mutation, these predictions indicate that all known *SSBP1* G-to-A mutations may be suitable candidates for base editing-based gene therapy in clinical applications. Notably, the most prevalent mutation reported to date (c.113G>A) is predicted to have the highest editing efficiency using a NG-Cas9 based ABE, which suggests a large percentage of patients could benefit from this strategy.

**Figure 5.**
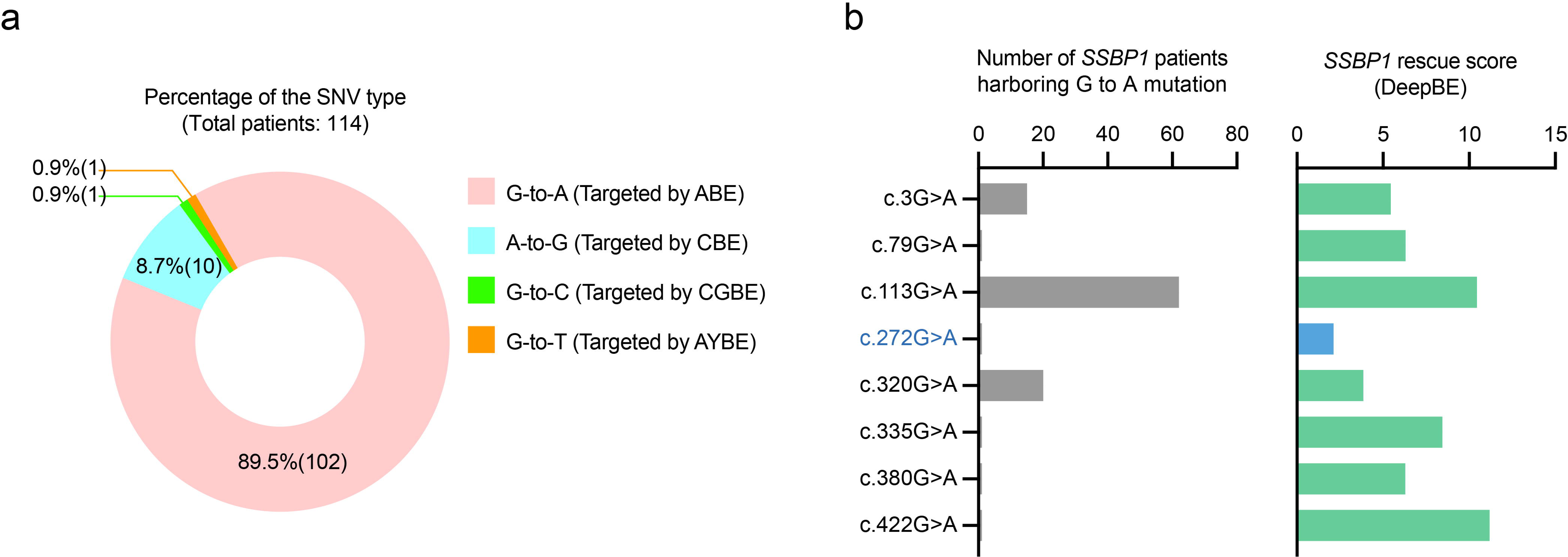
Characterizations of *SSBP1* mutations in patients and predictions of base editing efficacy. (a) Type and distribution of the 114 SNVs reported in the literature for *SSBP1*. Among them, 89.5% display G-to-A single base transitions, whereas 8.7% manifest A-to-G transitions, indicating the dominance of single base transition mutations. (b) For each *SSBP1* G-to-A disease-causing mutation, we assessed the number of patients and the corresponding *SSBP1* rescue score. We evaluated a total of 8 G-to-A mutations associated with mtDNA depletion syndrome for potential base editing applications. Predictions were carried out using the DeepBE software. The *SSBP1* rescue score indicates that the mutation is corrected without any bystander editing.

## Discussion

Mitochondrial replication plays an essential role in safeguarding the integrity and abundance of mtDNA to regulate cellular energy synthesis. Disruptions or anomalies in this complex process can lead to mitochondrial disorders that impair cellular vitality and function. Here, we report a novel mutation in *SSBP1* (c.272G>A:p.Arg91Gln) that impacted its multimer formation and DNA-binding affinity. These changes reduced mtDNA replication and altered mitochondria dynamics, which resulted in mtDNA depletion and mitochondrial dysfunction. Our findings replicate the from other reports of *SSBP1* mutations and mtDNA depletion-related mitochondrial disorders.

Mutations in *SSBP1* are associated with a variety of tissue-specific phenotypes, but primarily present as optic atrophy^15,16^. In vivo studies in *ssbp1*-null zebrafish mirrored the optic atrophy observed in patients with dominant *SSBP1* mutations^16^. Patients can also exhibit a spectrum of additional neurologic symptoms. Despite genotype-phenotype variabilities among different *SSBP1* mutations, reports of sensorineural deafness are particularly prevalent in the literature (**Supplementary** Figure 1). It is well known that clinical manifestations can overlap among various replisome components, including SSBP1, due to their interaction operative in the replisome and shared mechanisms in mtDNA replication and maintenance^11,12^. Sensorineural deafness is one of the overlapping phenotypes in mutations in mtDNA replisome machinery. However, the underlying mechanism of how hearing loss is linked to *SSBP1* mutations remains poorly understood. We observed that SSBP1 is expressed in the mouse cochlea, particularly its sensory hair cells, which requires substantial energy for mechanoelectrical sound transduction, ion recycling and homeostasis. Mitochondria regulate the function of hair cells and participate in apoptosis pathways that respond to intrinsic and extrinsic factors^55^. Thus, mtDNA depletion and subsequent mitochondrial dysfunction in cochlear hair cells may contribute to the development of sensorineural deafness in patients with *SSBP1* mutations.

To correct patient-derived cells harboring a heterozygous missense mutation (c.272G>A:p.Arg91Gln), we tested two ABE variants. The NG-ABE8e variant exhibited the highest editing efficacy and demonstrated optimal recovery of mitochondrial function. This can be attributed to the fact that most bystander editing results in silent mutations and that the off-target sites are not located within the open reading frame. The NG-ABE8eWQ had a more specific editing window and therefore displayed a diminished editing efficacy, but still showed substantial functional recovery. Importantly, although the mutations introduced into the NG-ABE8eWQ reduced overall editing capacity^56^, it also led to notably fewer off-target effects. The reason of diminished both on-target editing efficacy and undesired editing efficacy can be interpreted as the same tendency with ABE8eW invented by Liu’s group^56^. The ABE8e shows significantly high editing efficacy and a wide editing window which also induces a high frequency of undesired editing such as sgRNA-dependent off-target, RNA off-target, or bystander C editing. Therefore, Liu’s group applied TadA-V106W mutation to the ABE8e (ABE8eW) for decreasing RNA off-target^56^. Overall, the ABE8eW shows reduced RNA off-target editing efficacy but also relatively decreased on-target editing efficacy. In case of the ABE8eWQ, it shows further minimized RNA off-target editing and bystander C editing without any compromised on-target editing efficacy compared to the ABE8eW. From a perspective of functional restoration, the NG-ABE8e shows the best performance but, considering the safety issue, the NG-ABE8eWQ variant may be a better therapeutic candidate because it also shows meaningful functional restoration. As base editing makes its way into the clinic, our research offers evidence that base editing-based gene therapies are a promising treatment option for *SSBP1* mutations and other nuclear genomic mutations associated with mtDNA maintenance diseases.

Our findings indicate that correction of disease-causing pathogenic alleles via gene editing could facilitate the recovery of mitochondrial function. To our knowledge, this is the first report of the functional recovery of mitochondria diseases through base editing-based gene therapies. Base editor-treated patient-derived fibroblast displayed the improved mtDNA replication fidelity and the normalized mitochondrial morphology including the mitochondrial dynamics and ultrastructures. The morphological restoration of the mitochondria might be attributed to the increase of mtDNA abundance, modifications in mitochondrial dynamics related to fusion and fission, or a combination of both. Alternatively, base editing might moderate cellular senescence (**Supplementary** Figure 9), which possibly avert the mitochondrial dysfunction caused by *SSBP1* mutations.

In summary, we identified a novel *SSBP1* mutation and its causal relationship to mitochondrial disease. We also demonstrated gene editing could significantly enhance mtDNA replication fidelity and mtDNA copy number by correcting the mutation using two different base editors. Although base editing-based therapeutics have strong potential for mitochondrial diseases, substantial challenges persist before they can be clinically applied. Efficacy and specificity can be of particular concern in mitochondria diseases with multisystemic involvement. Nevertheless, several of the phenotypes-associated *SSBP1* mutations appear to be tissue-specific, primarily including optic atrophy and sensorineural deafness. The eye and ear sensory organs are strong candidates for gene therapies and genome editing due to their unique properties such as a small, enclosed compartment, immune privilege, and accessibility via established injection approach^57,58^. Therefore, we believe that base editing-based gene therapies may be a feasible strategy for correcting *SSBP1* mutations. In a clinical landscape where most patients with mitochondrial diseases lack a definitive treatment, the ability to restore mitochondrial function through base editing may be an attractive option for physicians and patients.

## Data Availability

All data produced in the present study are available upon reasonable request to the authors.

## Acknowledgements

This research was supported by the Korean Fund for Regenerative Medicine (KFRM) grant funded by the Korea government (the Ministry of Science and ICT, the Ministry of Health & Welfare (21A0202L1-11, Republic of Korea). This research was supported and funded by the SNUH Kun-hee Lee Child Cancer & Rare Disease Project, Republic of Korea (grant number: FP-2022-00001-004). This work was supported by the National Research Foundation of Korea (NRF) and funded by the Ministry of Education (grant number: 2022R1C1C1003147). This research was supported by the KAIST UP Program.

## Author Contributions

S.-Y.L. and S.B. designed research; J.H.C., S.-H.L., Y.Y., W.H.C., H.K., H.B.C., S.H.J., and J.P. contributed the experiments; S.-Y.L. contributed genomic analyses; S.-Y.L. and D.H.L. contributed the structural analysis; S.J.L., D.H.J., J.H.K., J.-J.S., J.-H.C., J.H.L., S.H.O. contributed reagents and analytic tools. S.-Y.L, S.B, and J.Y.K supervised and wrote the manuscript. All authors read and approved the final manuscript.

