## Supplementary materials for "Adenine base editing corrects point mutation in mitochondrial single-stranded binding protein (*SSBP1*) to improve mitochondrial function"

This PDF file includes:

- Supplementary Figure 1-10
- Supplementary Tables 1-4
- Supplementary Methods

**Supplementary Figure 1. Representation of genotypes and clinical phenotypes of patients with *SSBP1* mutations**

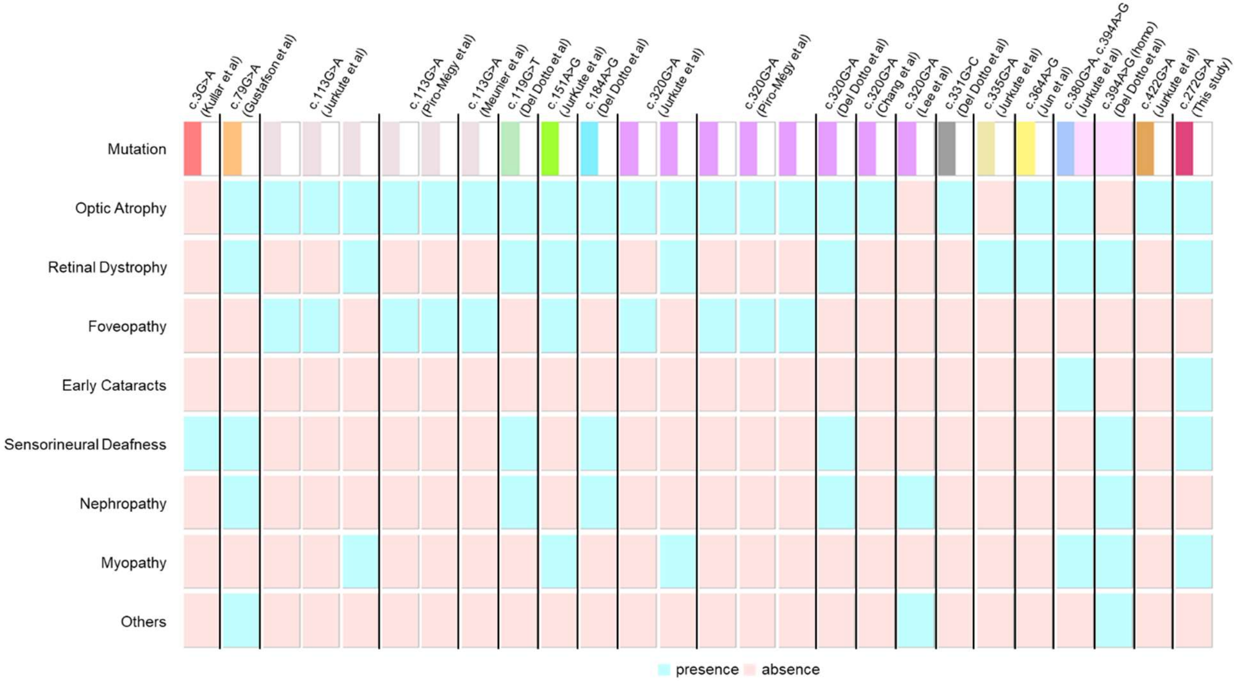


Our rare-grid plot comprises 14 mutations from 26 distinct families. Each column in this grid captures the genotype and clinical phenotype of an individual proband with a genetic completion. We conducted review of the clinical phenotypes documented in the literature, including optic atrophy, retinal dystrophy, foveopathy, early cataract, sensorineural deafness, nephropathy, myopathy, and other related conditions. In this illustration, blue squares indicate the presence of a particular phenotype, while red squares denote its absence. “Others” described in the rare-grid plot (y-axis) encompass anemia, bone marrow failure, ptosis, ophthalmoplegia, ataxia, metabolic strokes, multiple endocrine deficiencies, cardiomyopathy, pancytopenia, exocrine pancreatic insufficiency, adrenal cortical insufficiency, and developmental delay.

**Supplementary Figure 2. Immunohistochemical analysis of SSBP1 expression in the inner ear of P5 mice.**

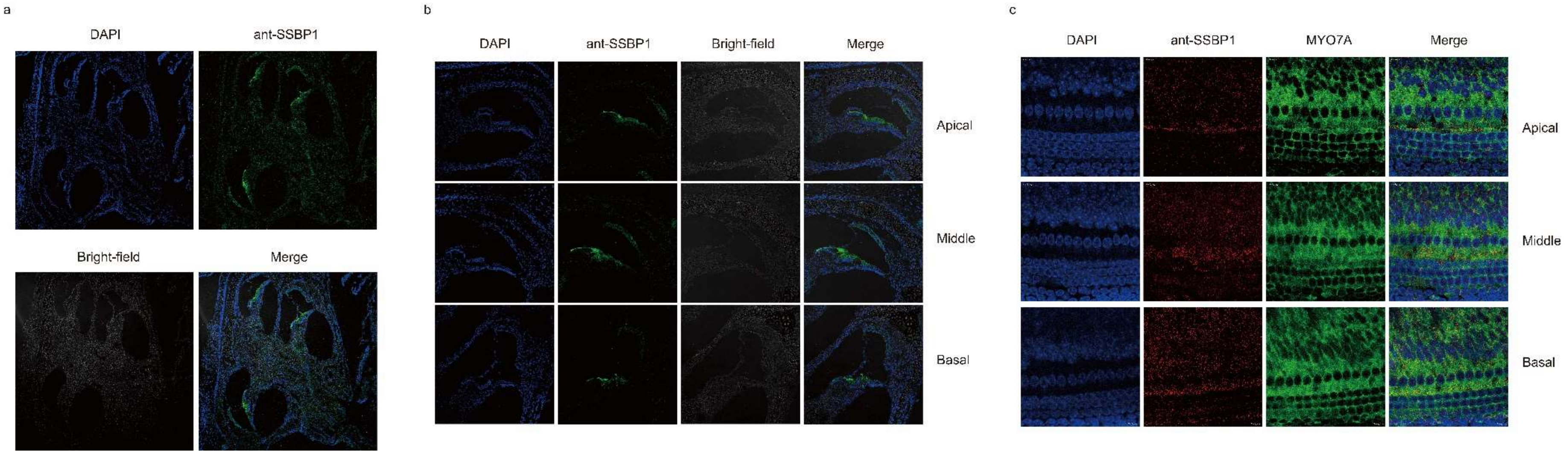


(a-b) Cochlear Overview: SSBP1 expression is ubiquitously observed throughout various structures in the cochlea. This includes the outer and inner hair cells of the organ of Corti, stria vascularis, spiral ligament, and spiral ganglion cells. The SSBP1 expression is notably expressed in hair cells of the organ of Corti and stria vascularis. (c) In the merged whole-mount image, there is a noticeable overlap between SSBP1 and Myosin VIIA, visible across the basal, middle, and apical turns. Myosin VIIA is stained by antibodies targeting cochlear hair cells. Moreover, SSBP1 is distinctly expressed in the supporting cells. The antibodies used are as follows: anti-SSBP1 (STJ95791-20, St John's Laboratory) and anti-Myo7A (sc-74516, Santa Cruz Biotechnology).

**Supplementary Figure 3. MLPA analysis to identify copy number variations in mitochondria genome isolated from patient's blood sample and fibroblast cells.**

(a)

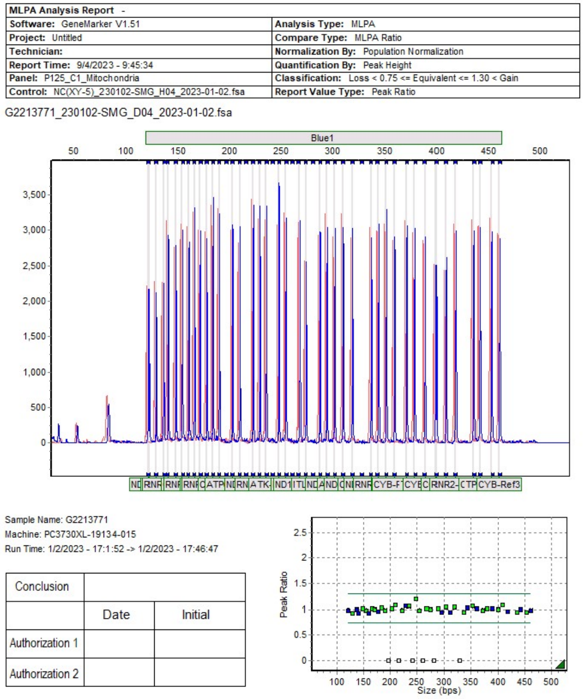


| | Probe Name | Bin Size | G2213771_230102-SMG_D0 |
|---|---|---|---|
| 1 | **ATP6-1** | 236.0 | 1.059 |
| 2 | **ATP6-2** | 302.0 | 1.045 |
| 3 | **ATP6-8993G** | 197.0 | -1 |
| 4 | **ATP6-8993T** | 203.0 | 1.021 |
| 5 | **CO1-1** | 148.0 | 1.009 |
| 6 | **CO1-2** | 400.0 | 0.997 |
| 7 | **CO1-3** | 436.0 | 0.947 |
| 8 | **CO2-1** | 166.0 | 1.020 |
| 9 | **CO2-2** | 184.0 | 1.032 |
| 10 | **CO2-3** | 210.0 | 1.081 |
| 11 | **CO3-1** | 319.0 | 1.048 |
| 12 | **CO3-2** | 352.0 | 1.067 |
| 13 | CYB-Ref1 | 360.0 | 1.016 |
| 14 | CYB-Ref2 | 389.0 | 1.016 |
| 15 | CYB-Ref3 | 461.0 | 0.976 |
| 16 | **ND1-3460A** | 261.0 | -1 |
| 17 | **ND1-3460G** | 267.0 | 1.009 |
| 18 | ND1-Ref | 137.0 | 0.997 |
| 19 | ND1-Ref2 | 295.0 | 0.941 |
| 20 | **ND2** | 454.0 | 0.939 |
| 21 | ND2-Ref | 310.0 | 0.941 |
| 22 | **ND3** | 380.0 | 1.018 |
| 23 | **ND4-1** | 155.0 | 0.972 |
| 24 | **ND4-11778A** | 216.0 | -1 |
| 25 | **ND4-11778G** | 223.0 | 0.977 |
| 26 | **ND4-2** | 172.0 | 0.995 |
| 27 | **ND4-3** | 409.0 | 1.074 |
| 28 | **ND5-1** | 129.0 | 0.929 |
| 29 | **ND5-2** | 190.0 | 0.977 |
| 30 | **ND5-3** | 255.0 | 0.979 |
| 31 | **ND5-4** | 274.0 | 0.996 |
| 32 | **ND6-14484C** | 328.0 | -1 |
| 33 | **ND6-14484T** | 337.0 | 0.953 |
| 34 | ND6-Ref | 122.0 | 0.980 |
| 35 | RNR1-Ref1 | 140.0 | 0.934 |
| 36 | RNR1-Ref2 | 160.0 | 0.929 |
| 37 | RNR1-Ref3 | 178.0 | 0.966 |
| 38 | RNR2-Ref1 | 229.0 | 1.060 |
| 39 | RNR2-Ref2 | 343.0 | 1.031 |
| 40 | RNR2-Ref3 | 418.0 | 0.966 |
| 41 | **TC** | 372.0 | 0.975 |
| 42 | **TK-8344A** | 248.0 | 1.194 |
| 43 | **TK-8344G** | 242.0 | -1 |
| 44 | **TL1-3243A** | 288.0 | 1.014 |
| 45 | **TL1-3243G** | 281.0 | -1 |
| 46 | TP-Ref | 442.0 | 0.993 |

(b)

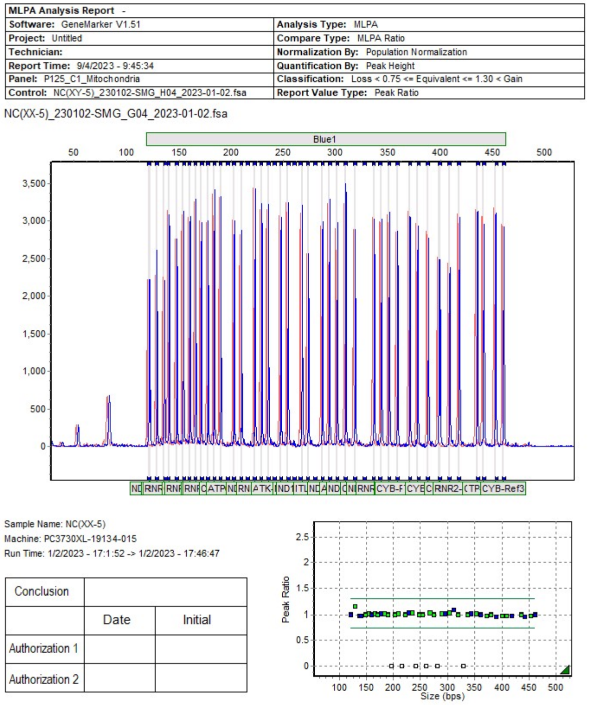


| | Probe Name | Bin Size | NC(XX-5)_230102-SMG_G0 |
|---|---|---|---|
| 1 | **ATP6-1** | 236.0 | 1.023 |
| 2 | **ATP6-2** | 302.0 | 1.028 |
| 3 | **ATP6-8993G** | 197.0 | -1 |
| 4 | **ATP6-8993T** | 203.0 | 0.998 |
| 5 | **CO1-1** | 148.0 | 1.000 |
| 6 | **CO1-2** | 400.0 | 0.986 |
| 7 | **CO1-3** | 436.0 | 0.991 |
| 8 | **CO2-1** | 166.0 | 1.011 |
| 9 | **CO2-2** | 184.0 | 1.020 |
| 10 | **CO2-3** | 210.0 | 1.018 |
| 11 | **CO3-1** | 319.0 | 0.999 |
| 12 | **CO3-2** | 352.0 | 1.011 |
| 13 | CYB-Ref1 | 360.0 | 1.005 |
| 14 | CYB-Ref2 | 389.0 | 0.970 |
| 15 | CYB-Ref3 | 461.0 | 0.991 |
| 16 | **ND1-3460A** | 261.0 | -1 |
| 17 | **ND1-3460G** | 267.0 | 1.032 |
| 18 | ND1-Ref | 137.0 | 0.978 |
| 19 | ND1-Ref2 | 295.0 | 1.016 |
| 20 | **ND2** | 454.0 | 0.980 |
| 21 | ND2-Ref | 310.0 | 1.084 |
| 22 | **ND3** | 380.0 | 0.990 |
| 23 | **ND4-1** | 155.0 | 1.015 |
| 24 | **ND4-11778A** | 216.0 | -1 |
| 25 | **ND4-11778G** | 223.0 | 0.998 |
| 26 | **ND4-2** | 172.0 | 0.991 |
| 27 | **ND4-3** | 409.0 | 0.977 |
| 28 | **ND5-1** | 129.0 | 1.144 |
| 29 | **ND5-2** | 190.0 | 1.004 |
| 30 | **ND5-3** | 255.0 | 1.000 |
| 31 | **ND5-4** | 274.0 | 1.000 |
| 32 | **ND6-14484C** | 328.0 | -1 |
| 33 | **ND6-14484T** | 337.0 | 0.996 |
| 34 | ND6-Ref | 122.0 | 0.999 |
| 35 | RNR1-Ref1 | 140.0 | 0.982 |
| 36 | RNR1-Ref2 | 160.0 | 1.001 |
| 37 | RNR1-Ref3 | 178.0 | 1.007 |
| 38 | RNR2-Ref1 | 229.0 | 1.022 |
| 39 | RNR2-Ref2 | 343.0 | 1.013 |
| 40 | RNR2-Ref3 | 418.0 | 0.987 |
| 41 | **TC** | 372.0 | 0.976 |
| 42 | **TK-8344A** | 248.0 | 0.992 |
| 43 | **TK-8344G** | 242.0 | -1 |
| 44 | **TL1-3243A** | 288.0 | 1.020 |
| 45 | **TL1-3243G** | 281.0 | -1 |
| 46 | TP-Ref | 442.0 | 0.967 |

(a) Absence of copy number variations in patient's blood sample.

(b) Absence of copy number variantions in patient's fibroblast cells.

**Supplementary Figure 4. Long-range PCR did not show any evidence of single mtDNA deletions in the patient.**

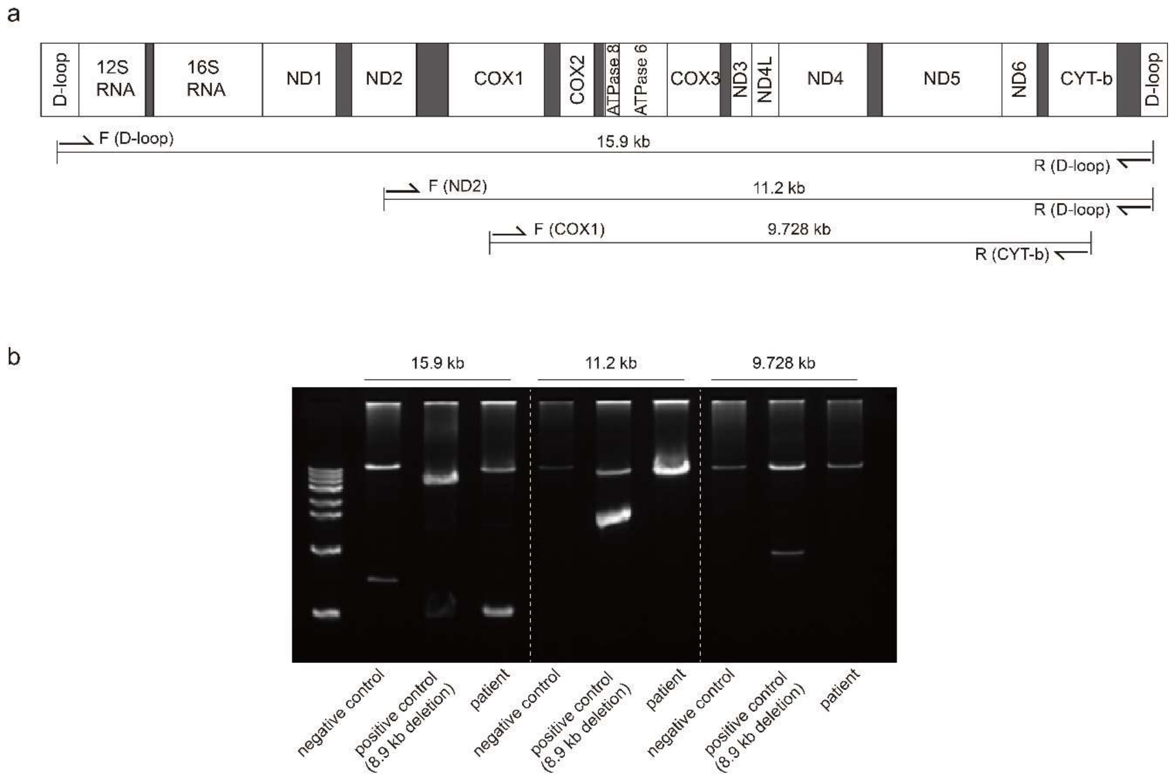


(a) Location of the mtDNA products amplified with our long-range PCR primer pairs: 15.9 kb (321-16271), 12.2 kb (5250-16271), 9.728 kb (5913-15661). (b) Long-range PCR was performed on genomic DNA obtained from three sources: a negative control without mtDNA deletions, a positive control harboring an 8.9 kb mtDNA deletion, and genomic DNA isolated from the proband's skeletal muscle.

**Supplementary Figure 5. Colocalization of wild-type and mutant SSBP1 with mitochondria in A549 cells**

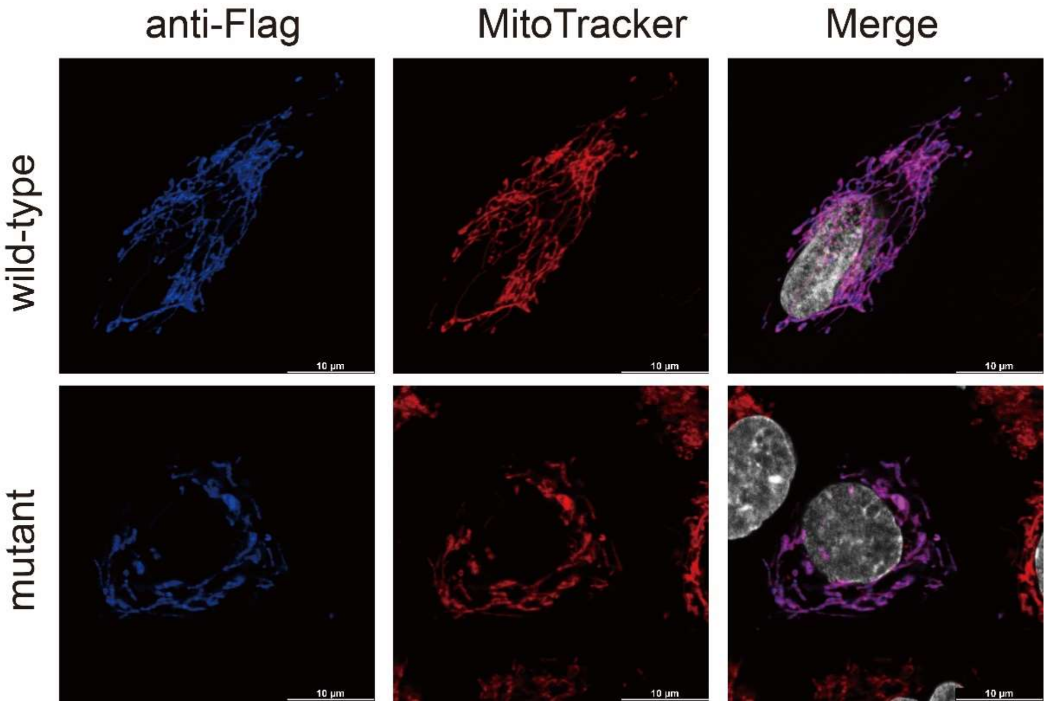


Both overexpressed wild-type and mutant SSBP1 (anti-Flag) showed colocalization with mitochondria. SSBP1 was visualized with Anti-Flag staining (blue), while mitochondria were highlighted using MitoTracker (red) and nuclei were stained with DAPI (white).

**Supplementary Figure 6. Analysis of SSBP1 binding to ssDNA using Electrophoretic Mobility Shift Assay (EMSA)**

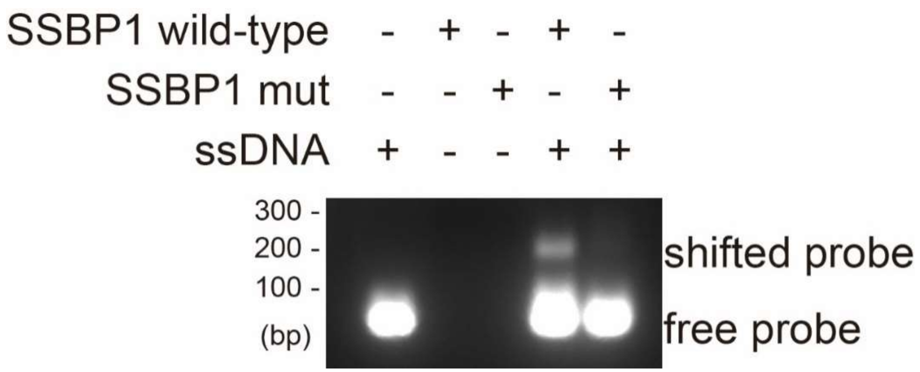


An Electrophoretic Mobility Shift Assay (EMSA) was used to analyze SSBP1. The recombinant proteins were incubated with single-stranded DNA (ssDNA). Following this, agarose gel electrophoresis confirmed the size shift of the ssDNA probe. Notably, only the wild-type SSBP1 clearly induced a shift in the probe.

**Supplementary Figure 7. Representative confocal images of Edu incorporation in A549 cells transfected with SSBP1 overexpression**

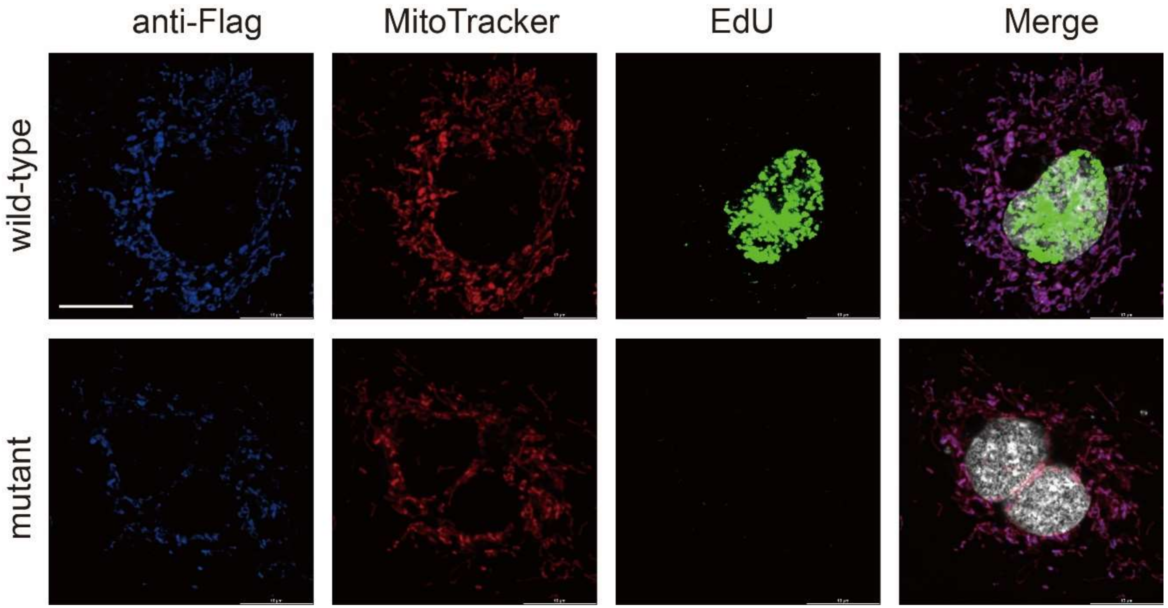


Representative images of EdU incorporation in A549 cells transiently overexpressed SSBP1 wild-type and mutant. In mutant cells, efficiency of mtDNA replication was inhibited as demonstrated by the Edu signal (green). SSBP1 was visualized with Anti-Flag staining (blue), while mitochondria were highlighted using MitoTracker (red).

**Supplementary Figure 8. OXPHOS complex expression levels in A549 cells overexpressing wild-type and mutant SSBP1**

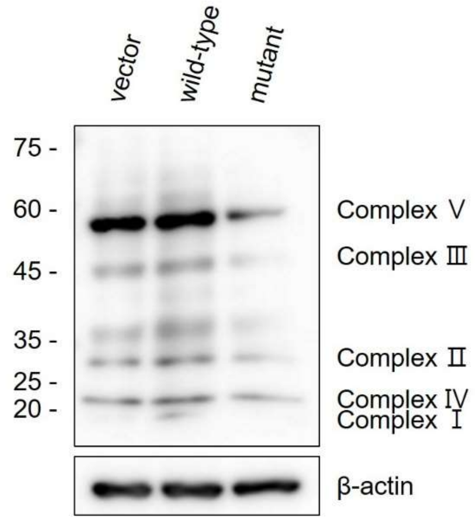


Immunoblot analysis was conducted to assess the expression levels of the oxidative phosphorylation system (OXPHOS) complex and β-actin in A549 cells overexpressing either wild-type or mutant SSBP1. In cells overexpressing the wild-type SSBP1, there was an upregulation of the OXPHOS complex relative to the vector-only controls. In contrast, cells with the mutant SSBP1 exhibited a downregulation of the OXPHOS complex.

**Supplementary Figure 9. Cellular senescence of patient and control fibroblasts depending on the cell passages**

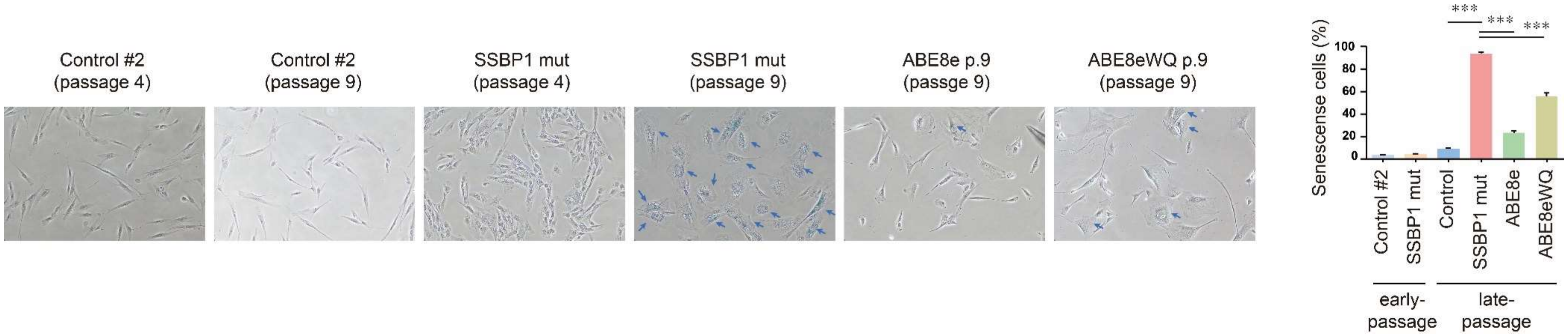


Senescence-associated β-galactosidase (SA-β-gal) was assessed with fibroblast cell lines at both early (passage 4) and late passages (passage 9). The blue arrow points to cells that are SA-β-gal-positive. Patient-derived fibroblasts displayed an expansive cytoplasm with numerous SA-β-gal-positive cells. The relative proportion of SA-β-gal positive cells in the total cell population was quantified. Data are presented as the means ± SEM (N=10-14). ***, $p<0.005$ (one-way ANOVA followed by the Bonferroni post hoc test).

**Supplementary Figure 10. SSBP1 protein expression levels in whole-cell lysates derived from fibroblast cell lines**

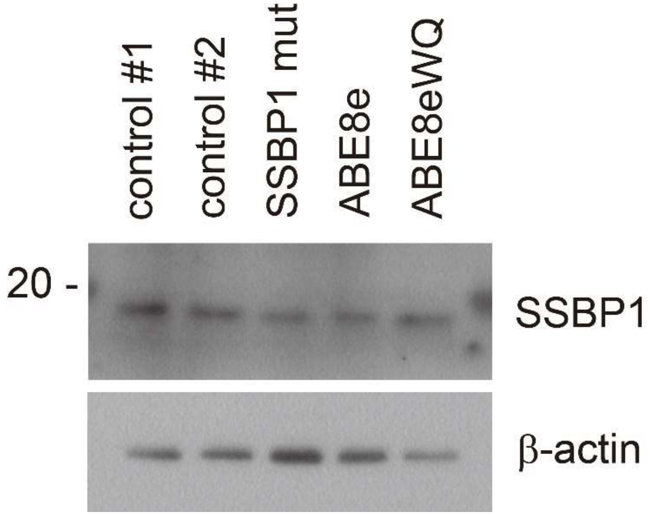


Fibroblast cell lines were lysed using RIPA lysis buffer to obtain whole-cell lysates. The extracted lysates were then separated using sodium dodecyl sulfate-polyacrylamide gel electrophoresis (SDS-PAGE). Following electrophoretic separation, the resolved proteins were transferred onto a suitable membrane and immunoblotted (IB) using the specified antibodies.

### Supplementary Table 1. Summary of reported *SSBP1* mutations, including this study, and its pathogenicity prediction analysis

| Genomic position (GRCh37/hg19) | HGVS | | Location (Exon/domain) | In-Silico Prediction | | | | Minor Allele Frequency | | Clinvar | ACMG/AMP guideline | |
|---|---|---|---|---|---|---|---|---|---|---|---|---|
| | Nucleotide change | Amino Acid change | | CADD phred | REVEL | SIFT | ClinPred | KOVA/KRGDB | gnomAD | Classification | Criteria | Classification |
| Chr7:141438969-G-A | c.3G>A | p.Met1? | Exon 2 / Absent | 23.0 | NA | 0.009 (Damaging) | 0.426 | ND/ND | exome(5.388e-05) gnome(3.186e-05) | ND | PM2, PP1,PP4, PS3_moderate | Likely Pathogenic |
| Chr7:141442023-G-A | c.79G>A | p.Glu27Lys | Exon 3 / Absent | 32.0 | 0.319 | 0.005 (Damaging) | 0.949 (Damaging) | ND/ND | ND | ND | PS2,PM2,PP4, PS3_moderate | Likely Pathogenic |
| Chr7:141443388-G-A | c.113G>A | p.Arg38Gln | Exon 4 / SSB | 32.0 | 0.514 | 0.003 (Damaging) | 0.974 (Damaging) | ND/ND | ND | Pathogenic | PS1,PS3,PM2,PP1,PP4 | Pathogenic |
| Chr7:141443394-G-T | c.119G>T | p.Gly40Val | Exon 4 / SSB | 12.59 | 0.815 | 0 (Damaging) | 0.998 (Damaging) | ND/ND | ND | Likely Pathogenic | PS2,PS3,PM2,PP3,PP4 | Pathogenic |
| Chr7:141443426-A-G | c.151A>G | p.Lys51Glu | Exon 4 / SSB | 23.8 | 0.494 | 0.996 | 0.775 (Damaging) | ND/ND | ND | ND | PM2, PP1, PP4 | Uncertain Significance |
| Chr7:141443459-A-G | c.184A>G | p.Asn62Asp | Exon 4 / SSB | 23.1 | 0.21 | 0.323 | 0.846 (Damaging) | ND/ND | ND | ND | PS2, PS3, PM2, PP1, PP4 | Pathogenic |
| Chr7:141443747-G-A | c.272G>A | p.Arg91Gln | Exon 5 / SSB | 23.9 | 0.176 | 0.032 (Damaging) | 0.924 (Damaging) | ND/ND | ND | ND | PS2, PS3, PM2, PP4 | Pathogenic |
| Chr7:141445301-G-A | c.320G>A | p.Arg107Gln | Exon 6 / SSB | 25.0 | 0.244 | 0 (Damaging) | 0.983 (Damaging) | ND/ND | ND | Pathogenic | PS1, PS2, PS3, PM2, PP1, PP4 | Pathogenic |
| Chr7:141445312-G-C | c.331G>C | p.Glu111Gln | Exon 6 / SSB | 25.8 | 0.32 | 0.368 | 0.951 (Damaging) | ND/ND | ND | ND | PS3, PM2, PP4 | Likely Pathogenic |
| Chr7:141445316-G-A | c.335G>A | p.Gly112Glu | Exon 6 / SSB | 28.6 | 0.68 | 0 (Damaging) | 1 (Damaging) | ND/ND | ND | ND | PS2, PM2, PP4 | Likely Pathogenic |
| Chr7:141445345-A-G | c.364A>G | p.Lys122Glu | Exon 6 / SSB | 24.8 | 0.696 | 0.35 | 0.843 (Damaging) | ND/ND | ND | ND | PS3, PM2, PP1, PP4 | Likely Pathogenic |
| Chr7:141445361-G-A | c.380G>A | p.Arg127Gln | Exon 6 / SSB | 25.7 | 0.688 | 0.039 (Damaging) | 0.270 (Damaging) | ND/ND | exome(3.989e-05) gnome(ND) | ND | PM2, PP1, PP4 | Uncertain Significance |
| Chr7:141445375-A-G | c.394A>G | p.Ile132Val | Exon 6 / SSB | 20.6 | 0.113 | 0.207 | 0.546 (Damaging) | ND/ND | exome(1.196e-05) gnome(3.184e-05) | Uncertain Significance | PS1, PM2, PP1, PP4 | Likely Pathogenic |

| | | | | | | | | | | | | |
|---|---|---|---|---|---|---|---|---|---|---|---|---|
| Chr7:14145 0129-G-A | c.422G>A | p.Ser141Asn | Exon 7 / SSB | 22.5 | 0.084 | 0.2 | 0.869 (Damaging) | ND/ND | ND | Pathogenic | PS2, PS3, PM2, PP4 | Pathogenic |

Abbreviations: SSB, single-stranded binding domain; ND, not determined; NA, not available

Refseq transcript accession number NM_003143.3; Refseq protein accession number NP_003143.1

HGVS: Human Genome Variation Society (https://www.hgvs.org/)

CADD: Combined Annotation Dependent Depletion (https://cadd.gs.washington.edu/ )

REVEL: Rare Exome Variant Ensemble Learner (https://sites.google.com/site/revelgenomics/)

SIFT: Sorting Intolerant From Tolerant (https://sift.bii.a-star.edu.sg/)

ClinPred: Prediction tool to identify disease-relevant nonsynonymous single nucleotide variants (https://sites.google.com/site/clinpred/)

KOVA: Korean Variant Archive (https://www.kobic.re.kr/kova/)

KRGDB: Korean Reference Genome Database (http://coda.nih.go.kr/coda/KRGDB/index.jsp)

gnomAD: The Genome Aggregation Database (https://gnomad.broadinstitute.org/)

ACMG/AMP 2015 guideline (http://wintervar.wglab.org/)

* Analysis of ACMG/AMP guidelines for conflicting or uncertain significance variants in the Clinvar database

**Supplementary Table 2. Summary of reported genotypes and phenotypes associated with *SSBP1* mutations.**

| Reference | Family (number) | Affected patients (number) | Gene | Mutation | Zygosity | mtDNA depletion | mtDNA deletion | Clinical phenotypes |
|---|---|---|---|---|---|---|---|---|
| Kullar et al | 1 | 15 | *SSBP1* | NM_001256510:c.3G>A | heterozygote | O | O | Sensorineural deafness |
| | | | *MT-RNR1* | m.1555A>G | homoplasmy | | | |
| Jurkute et al | 2 | 5 | *SSBP1* | NM_001256510.1:c.113G>A | heterozygote | NA | NA | Optic atrophy, Foveopathy |
| | 1 | 12 | *SSBP1* | NM_001256510.1:c.320G>A | heterozygote | NA | NA | Optic atrophy, Foveopathy |
| | 1 | 1 | *SSBP1* | NM_001256510.1:c.422G>A | heterozygote | NA | NA | Optic atrophy |
| Piro-Mégy et al | 2 | 29 | *SSBP1* | NM_003143.3:c.113G>A | heterozygote | O | X | Optic atrophy, sometimes foveopathy |
| | 3 | 3 | *SSBP1* | NM_003143.3:c.320G>A | heterozygote | O | X | Optic atrophy, sometimes foveopathy |
| Gustafson et al | 1 | 1 | *SSBP1* | NM_003143.3:c.79G>A | heterozygote | O | O | Infantile anemia, bone marrow failure, growth failure, ptosis, ophthalmoplegia, ataxia, retinal dystrophy, Sensorineural deafness, kidney disease, metabolic strokes, multiple endocrine deficiencies |
| | | | | m.8629_14068del5440 | | | | |
| Del Dotto et al | 1 | 1 | *SSBP1* | NM_003143.3:c.119G>T | heterozygote | O | X | optic atrophy, retinal degeneration, nephropathy, Sensorineural deafness |
| | 1 | 3 | *SSBP1* | NM_003143.3:c.184A>G | heterozygote | O | X | optic atrophy, retinal macular dystrophy, Sensorineural deafness, nephropathy |

| | | | | | | | | |
|---|---|---|---|---|---|---|---|---|
| | 1 | 2 | *SSBP1* | NM_003143.3:c.320G>A | heterozygote | O | X | optic atrophy, retinal macular dystrophy, Sensorineural deafness, nephropathy |
| | 1 | 1 | *SSBP1* | NM_003143.3:c.331G>C | heterozygote | O | X | optic atrophy |
| | 1 | 1 | *SSBP1* | NM_003143.3:c.394A>G | homozygote | O | X | retinal dystrophy, Sensorineural deafness, cardiomyopathy, ataxia, nephropathy, growth retardation |
| Jurkute et al | 1 | 3 | *SSBP1* | NM_003143:c.151A>G | heterozygote | NA | NA | Myopia, Optic atrophy, Retinal dystrophy, Foveopathy, Attenuated retinal vessels |
| | 1 | 1 | *SSBP1* | NM_003143:c.113G>A | heterozygote | NA | NA | Optic atrophy, Retinal dystrophy, Attenuated retinal vessels, Myopia |
| | 1 | 1 | *SSBP1* | NM_003143:c.320G>A | heterozygote | NA | NA | Optic atrophy, Retinal dystrophy, Attenuated retinal vessels, Myopia |
| | 1 | 1 | *SSBP1* | NM_003143:c.380G>A | compound heterozygote | NA | NA | Optic atrophy, Retinal dystrophy, Attenuated retinal vessels, Early cataracts, Myopia |
| | | | *SSBP1* | NM_003143:c.394A>G | | NA | NA | |
| | 1 | 1 | *SSBP1* | NM_003143:c.335G>A | heterozygote | NA | NA | Retinal dystrophy |
| Meunier et al | 1 | 27 | *SSBP1* | NM_003143.3:c.113G>A | heterozygote | X | X | optic atrophy, visual impairment, foveopathy |
| Lee et al | 1 | 1 | *POLG* | NM_002693.3_c.868C>T | heterozygote | N/A | O | sideroblastic anemia, pancytopenia, bone marrow failure, proximal renal tubular acidosis, |
| | | | *SSBP1* | NM_003143.3:c.320G>A | heterozygote | | | |

| | | | | | | | | |
|---|---|---|---|---|---|---|---|---|
| | | | | | | | | hronic kidney disease, exocrine pancreatic insufficiency, adrenal cortical insufficiency, developmental delay |
| Jun et al | 1 | 2 | *SSBP1* | NM_003143.3:c.364A>G | heterozygote | O | X | optic atrophy, diffuse retinal nerve fiber layer thinning |
| Chang et al | 1 | 1 | *SSBP1* | NM_003143.3:c.320G>A | heterozygote | NA | NA | Optic atrophy, color blindness |
| This study | 1 | 1 | *SSBP1* | NM_003143.3:c.272G>A | heterozygote | O | X | Sensorineural deafness, optic atrophy, myopathy, early cataract, macular dystrophy |

Abbreviations: O, presence; X, absence; NA, not applicable.

### Supplementary Table 3. Oligonucleotides sequences used in this study

| |
|---|
| **Human *SSBP1* primer pair (cDNA)**<br>Forward : 5'-AAGATCCCTGAATCGTGTGC-3'<br>Reverse : 5'-TCGAGACCCCTTTTTCACAT-3' |
| **Human *7s DNA* primer pair (gDNA)**<br>Forward : 5'-GTGGCTTTGGAGTTGCAGTT-3'<br>Reverse 1 (7s + D-Loop) : 5'-CAGCCACCATGAATATTGTAC-3'<br>Reverse 2 (7s + D-Loop) : 5'-GAAGCAGATTTGGGTACCAC-3' |
| **Human *ND1* primer pair (gDNA)**<br>Forward : 5'-TACGGGCTACTACAACCCTTC-3'<br>Reverse : 5'-ATGGTAGATGTGGCGGGTTT-3' |
| **Human *ND5* primer pair (gDNA)**<br>Forward : 5'-CATTACTAACAACATTTCCCCCGC-3'<br>Reverse : 5'-GGCTGTGAGTTTTAGGTAGAGGG-3' |
| **Human *SLCO2B1* primer pair (gDNA)**<br>Forward : 5'-CCTGATGCCTAGGTTTCTTTTCTTG-3'<br>Reverse : 5'-GGTCATCTGCCTACCCTAGAAC-3' |
| **Human *SERP1NA1* primer pair (gDNA)**<br>Forward : 5'-CAGTGAATAAATGAGGCGTACATCC-3'<br>Reverse : 5'-GACTGTTTCTCATGCCTCTGGAAAG-3' |
| **Human *SSBP1* primer pair-probe set (gDNA)**<br>Forward : 5'-AGGTGATGTCAGTCAAAAGA-3'<br>Probe For WT: HEX- ATCAGTATTCCGGCCAGGCCTCA -BHQ1 |

| |
|---|
| Probe For MUT: FAM- ATCAGTATTCCAGCCAGGCCTCA -BHQ1<br>Reverse : 5'-CCATGAGAACACTTCTTATCG-3' |
| **Long-range PCR 1.59 kb amplicon primer pair (gDNA)**<br>D-loop Forward : 5'-TGGCCACAGCACTTAAACACATCTC-3'<br>D-loop Reverse : 5'-GGAGTTGCAGTTGATGTGTG-3' |
| **Long-range PCR 11.2 kb amplicon primer pair (gDNA)**<br>ND2 Forward : 5'-TTGCCCAAATGGGCCATTAT-3'<br>D-loop Reverse : 5'-GGAGTTGCAGTTGATGTGTG-3' |
| **Long-range PCR 9.728 kb amplicon primer pair (gDNA)**<br>COX1 Forward : 5'-GACCGTTGACTATTCTCTAC-3'<br>CYTB Reverse : 5'-GGATGGATAGTAATAGGGCA-3' |
| **Human control siRNA for sense**<br>5'-CCUCGUGCCGUUCCAUCAGGUAGUU-3' |
| **Human control siRNA for anti-sense**<br>5'-CUACCUGAUGGAACGGCACGAGGUU-3' |
| **Human *SSBP1* siRNA #1 for sense**<br>5'- CAACAAUCAUAGCUGAUAAUAUU-3' |
| **Human *SSBP1* siRNA #1 for anti-sense**<br>5'-UAUUAUCAGCUAUGAUUGUUGUU-3' |
| **Human *SSBP1* siRNA #2 for sense**<br>5'-UAAUACAGGUCUUCGAAACAUUU-3' |
| **Human *SSBP1* siRNA #2 for anti-sense**<br>5'-AUGUUUCGAAGACCUGUAUUAUU-3' |
| **Electrophoretic mobility shift assay probe**<br>5'-GGACTATTTATTCAATATATTTAAGAACTAATTCCAGCTGAGCGCCGG-3' |

### Supplementary Table 4. Antibodies used in this study

| Name | Company | cat.no |
|---|---|---|
| anti-DNA mouse monoclonal | PROGEN | AC-30-10 |
| MitoTracker™ Red CMXRos | Invitrogen | M7512 |
| Anti-SSBP1 antibody | St John's Laboratory | STJ95791 |
| SSBP1 Polyclonal antibody | proteintech | 12212-1-AP |
| Monoclonal Anti-Flag M2 antibody produced in mouse | Sigma | F3165-2MG |
| Total OXPHOS Human WB antibody cocktail | abcam | ab110411 |
| Anti-beta Actin antibody (AC-15) | abcam | ab6276 |
| Anti-c-Myc (phospho S62) antibody | abcam | ab51156 |
| Goat anti-Mouse IgG(H+L)-HRP | GenDEPOT | SA001-500 |
| Goat anti-Rabbit IgG(H+L)-HRP | GenDEPOT | SA002-500 |
| Goat Anti-Rabbit IgG H&L (Alexa Fluor 488) | abcam | ab150077 |
| Goat Anti-Rabbit IgG H&L (Alexa Fluor 555) | abcam | ab150078 |
| Goat Anti-Mouse IgG H&L (Alexa Fluor 488) | abcam | ab150113 |
| Goat Anti-Mouse IgG H&L (Alexa Fluor 647) | abcam | ab150115 |

### Supplementary Methods

#### Molecular genetic testing

In the initial phase (Step 1), we examined 22 mutations across 10 established deafness genes (*GJB2*, *SLC26A4*, *TMPRSS3*, *CDH23*, *OTOF*, *TMC1*, *ATP1A3*, *MPZL2*, *COCH*, and *12S rRNA*) through genotyping techniques such as custom capillary sequencing and U-TOP™ HL Genotyping Kits[1,2]. During the subsequent phase (Step 2), targeted sequencing was achieved using whole-exome sequencing (WES). For this, target regions were captured employing the SureSelectXT Human All Exon V5 kit for WES (Agilent Technologies, Santa Clara, CA, USA). Libraries were constructed in line with the manufacturer's protocol and subjected to paired-end sequencing on the NovaSeq 6000 system (Illumina, San Diego, CA, USA). In the concluding phase (Step 3), patients not yet diagnosed underwent WGS followed by comprehensive bioinformatic analysis and curation. DNA libraries were assembled using the TruSeq DNA PCR-Free Library Prep Kits (Illumina) and sequenced on the Illumina NovaSeq6000 platform with an average coverage depth of 30×. Resultant genome sequences were mapped to the human reference genome (GRCh38) utilizing the BWA-MEM algorithm. PCR duplicates were eliminated with SAMBLASTER. Primary mutation detection for base substitutions and short indels was conducted using HaplotypeCaller2 and Strelka2, correspondingly. Subsequently, mutations underwent sieving, Mendelian inheritance patterns were assessed, and potential *de novo* mutations, along with their predicted impacts, were identified. The parental samples were used for Sanger sequencing to determine the phasing. Medical geneticists made the final assessment of mutation pathogenicity, according to the ACMG-AMP guideline[3].

**mtDNA panel sequencing and MLPA**

For mitochondria panel sequencing, DNA was extracted from peripheral blood samples using the Chemagic 360 instrument (Perkin Elmer, Baesweiler, Germany). The complete human mitochondrial genome was amplified in two overlapping fragments: fragment I (spanning 9,289 bp), and fragment II (spanning 7,626 bp). Fragment 1 was amplified using the primer pair hmtF1 569 (5′-AACCAAACCCCAAAGACACC-3′) and hmtR1 9819 (5′-GCCAATAATGACGTGAAGTCC-3′), and fragment II was amplified using the primer pair htmF2 9611 (5′-TCCCACTCCTAAACACATCC-3′) and hmtR2 626 (5′- TTTATGGGGTGATGTGAGCC-3′). PCR reactions were conducted using the following cycling parameters: initial denaturation at 94 °C for 2 min; 10 cycles of 94 °C for 15 s, 65 °C for 30 s, and 68 °C for 5 min; 25 cycles of 94 °C for 15 s, 65 °C for 30 s, and 68 °C for 5 min; and a final extension at 68 °C for 7 min. Subsequently, a library was generated using the Nextera DNA Flex Library Prep Kit (Illumina) following the manufacturer’s instructions. Paired-end sequencing was performed with generation of 150-bp reads on the MiSeq platform (Illumina). Bioinformatic processes, including alignment and annotation, were performed using NextGene Version 2.4.0.1 (Softgenetics, State College, PA, USA). Furthermore, we performed the SALSA MLPA probemix P125-C1) (Lot#0719) (MRC-Holland, Amsterdam, the Netherlands) to detect the deletions using 32 Probes. We analyzed the amplification products using an ABI PRISM 3130 Genetic Analyzer (Applied Biosystems, Foster City, CA) and interpreted the results using Gene Marker 1.91 software (SoftGenetics, State College, PA).

**SDS-PAGE and immunoblotting**

Cultured cells were washed twice with PBS, and whole-cell lysates were prepared in RIPA buffer supplemented with a protease inhibitor cocktail. For SDS-PAGE, lysates mixed with NuPAGE 4X LDS sample buffer (NP0007, Invitrogen) and denatured at 85°C for 10 min. separated proteins by SDS-PAGE were transferred to polyvinylidene difluoride (PVDF) membrane. The membranes were blocked with 5% nonfat milk in TBS-T solution and incubated with the primary antibodies. The membranes were then washed with TBS-T solution several times and followed by a horseradish peroxidase-conjugated anti-rabbit IgG or anti-mouse IgG antibody. After antibody incubation, protein band were detected by chemiluminescence reagent (RPN2106, Cytiva). Antibodies used in this study are summarized in Supplementary Table 4.

**Purification of recombinant SSBP1 proteins**

To purify the wild-type and its mutants, *E. coli* strain BL21 (DE3) cells (Agilent) were cultured at 37 °C until the OD at 600 nm reached 0.6. Subsequently, 0.5 mM isopropyl-β-d-thiogalactopyranoside (IPTG) was introduced to induce SSBP1 expression, and cells were allowed to grow 16 hours at 16 °C. Upon completion of expression, the cells were collected by centrifugation and lysed using sonication in a lysis buffer (50 mM Tris buffer pH 7 containing 250 mM NaCl, 4 mM MgCl2, 25 μg/mL DNase I, 25 μg/mL RNase A, and a protease inhibitor cocktail). The resulting supernatant was applied to a 5 mL HiTrap IMAC FastFlow column (GE

Healthcare Life Sciences) utilizing the ÄKTA FPLC system (GE Healthcare Life Sciences). The column was washed with five column volumes of washing buffer (50 mm Tris pH 7.0 with 250 mM NaCl), and elution was carried out using an imidazole step gradient. After elution, the proteins were subjected to dialysis, followed by size exclusion chromatography (SEC). These dialyzed proteins were loaded onto a HiLoad 16/600 Superdex 200 pg by fast protein liquid chromatography. To each fraction, 10% glycerol was added, and the proteins were stored at −80 °C.

**Electrophoretic mobility shift assay**

The electrophoretic mobility shift assay (EMSA) for SSBP1 was conducted following previously established methods[4,5]. Briefly, we utilized custom-synthesized ssDNA oligonucleotides (Supplementary Table 3), provided by Cosmo Genetech (Korea). In the experimental procedure, varying concentrations of both wild-type and mutant SSBP1 recombinant proteins were incubated with a known quantity of single-stranded oligonucleotide probes in a binding buffer (composed of 50 mM Tris pH 7.5, 10% glycerol, and 150 mM NaCl) for a duration of 30 minutes at room temperature. Subsequently, a loading buffer (Dyne LoadingSTAR, DYNE BIO, Korea) was added, and the prepared samples were loaded onto 2% agarose gels. Electrophoresis was carried out in 1×Tris-borate-EDTA (TBE) buffer for 45 minutes at 100 V. Following electrophoresis, the gels were visualized using UV light.

**Immunocytochemistry and EdU labeling assay**

For immunofluorescence microscopy, cultured A549 cells on a cover glass were transfected with 1–2 μg of total plasmid DNA for 24 h. The cells were fixed with 4% paraformaldehyde, then washed with 0.1% Tween20 in PBS. Next permeabilized with 0.5% Triton X-100, 1% BSA in PBS. And then the cells were incubated with the primary antibody and Alexa Fluor-conjugated secondary antibody. The cells were mounted with 4′,6′-diamidino-2-phenylindole (DAPI)-containing mounting medium (ab104139, abcam). Confocal images were captured by a laser scanning confocal microscope (Leica STELLARIS 8, Upright). For EdU labeling (C10337, Invitrogen), EdU was added to the culture media at a concentration of 100 μM and incubated for 90–120 minutes. Subsequently, Alexa Fluor 488 azide was used for detection. After EdU labeling, cells were treated with MitoTracker Red (Invitrogen, M7512) to stain the mitochondria. For quantification, EdU foci that were positive for MitoTracker Red were counted in over 10 individual images for each cell line.

**RNA interference**

The siRNAs were synthesized by Genolution. siRNA duplexes (siRNA A and B) were transfected into A549 cells using RNAiMAX (13778075, Invitrogen) dissolved in Opti-MEM (11058021, Gibco), following the manufacturer’s protocols. The siRNA duplex used in this study are summarized in Supplementary Table 3. The final concentration of siRNA for transfection was 20 mM. After 24 h, cells were trypsinized, suspended in the transfection mixture, and re-plated. On the following day, cells were transfected again with the siRNA duplexes and grown for an additional 4 h for an effective knockdown.

**Oxygen consumption rate**

The intact cellular oxygen consumption rate (OCR) and extracellular acidification rate (ECAR) were measured in real time using the Seahorse XF96 Extracellular Flux Analyser (Seahorse Bioscience, North Billerica, MA, USA according to manufacturer's protocol. Briefly, 8.0 ×10^3 of fibroblasts or A549 cells were seeded into 96 well Seahorse microplates in 100 μL of growth medium and incubated at 37 ºC in 5% CO2 for 24 h and the calibrator plate was equilibrated in a non-CO2 incubator overnight. Before starting the test, cells were washed twice with assay running media (unbuffered DMEM, 25 mM glucose, 1 mM glutamine, 1 mM sodium pyruvate) and equilibrated in a non-$CO_2$ incubator. Once the probe calibration was completed, the probe plate was replaced by the cell plate. The protocol was optimized and gave the measurement of oxygen consumption rate (OCR) and extracellular acidification rate (ECAR) simultaneously. Totally, the assay protocol incorporated four compounds injection which could be applied to modulate mitochondrial function to determine the mitochondrial parameters, including basal respiration, maximal respiration, and ATP production. The analyzer plotted the value of OCR and the corresponding ECAR followed by injection of the compounds sequentially as follows: oligomycin (1 μM), an inhibitor of ATP synthase which leads to exhibit maximal glycolysis metabolism; followed by exposure of carbonycyanide p-(trifluoromethoxy) phenylhydrazone (FCCP) (1 μM), the uncoupler of ETC and OXPHOS which induces the peak oxygen consumption to evaluate the oxidative metabolism indirectly; by addition of ETC inhibitor rotenone and the complex III inhibitor antimycin A to uncover the part of non-mitochondrial respiration at a final concentration of 1 μM .

**Fibroblast cell culture**

The fibroblast culture was conducted following the methods described by Vangipuram, M., et al[6]. Under local anesthesia, a 4 mm round skin biopsy was collected from the donor's designated area and immediately placed in a 1 mL tube with 1 mL of Phosphate Buffered Saline (PBS). The biopsy was then transferred to a sterile 6-cm culture dish using sterile forceps and dissected into 9-12 uniformly sized pieces with sterile scalpel blades or scissors. Using pointed sterile forceps, each biopsy piece was placed into individual wells of a 12-well plate containing 200 µL of complete DMEM supplemented with 20% Fetal Bovine Serum (FBS), ensuring proper attachment to the bottom of the well. The plate was incubated at 37°C and monitored daily for the first week, with up to 50 µL of media added every 2 days to compensate for evaporation. When the fibroblasts formed a confluent monolayer, they were harvested for further expansion.

**Transmission electron microscopy**

Cells were fixed in a 2.5% Glutaraldehyde in PBS buffer 48 hour at 4°C and washed in PBS buffer 10 min. After that post-fixed in a 1% osmium tetroxide for 2 hours at room temperature. Following incubation, washed 2 times in PBS buffer. After that, treat ethanol solutions (30%-100%) for dehydration and embedded in EmBed 812. Ultrathin sections were used for making sample blocks. Staining the sections with uranyl acetate and lead citrate and examined in a JEOL, JEM1400 Flash transmission electron microscope at 80 kV.

**Senescence assay**

Patient-derived fibroblasts and edited cells were seeded in 6-well plates. After a 16-hour incubation period, the cells were carefully rinsed with PBS and subsequently fixed with a 1× fixative solution provided by a senescence β-galactosidase staining kit (9860, Cell Signaling Technology). The fixation process was carried out for 20 minutes at room temperature. A fresh β-galactosidase staining solution was meticulously prepared in accordance with the manufacturer's instructions. Following two PBS washes, each well was treated with 1 mL of the staining solution. Cells exhibiting β-galactosidase positivity were identified as senescent cells, and their population was quantified by analyzing a minimum of >500 cells. All images in each figure were captured using identical microscope settings and are representative of the entire cell population.
